# A calibration pipeline for controlled ocular photostimulation with consumer see-through extended-reality glasses

**DOI:** 10.64898/2026.08.26.26361398

**Authors:** Matt Gaidica, Matthew Rosengart

## Abstract

Consumer extended-reality (XR) glasses could provide controlled ocular light exposure, but their settings do not specify the light delivered at the eye. We developed and validated a calibration pipeline linking display control, corneal-plane spectroradiometry, settings-to-exposure modeling, and rendered application content for a see-through binocular micro-OLED display (VITURE Luma Ultra). Automated measurements characterized color channels, digital codes, hardware brightness levels, and duty-cycle settings across repeated fittings on a phantom head. The blue primary peaked at 459 nm with no resolved spectral shift across intensity settings. Blue melanopic equivalent daylight illuminance (EDI) ranged from 14.7 to 252 lx across hardware brightness levels, while full white delivered 269 lx melanopic EDI at its highest usable setting, exceeding the 250 lx daytime recommendation for healthy adults. A model calibrated on separate sessions predicted melanopic EDI for held-out codes, brightness levels, and color mixtures with a mean absolute percentage error of 3.4% (4.5% for photopic illuminance), allowing exposure to be estimated from logged display settings without remeasuring each uniform stimulus. Rendered application frames revealed position- and illuminated-area-dependent exposure; a spatial correction derived from an independent display grid accounted for these effects and closely reproduced measured frame exposure. Together, these results establish a reusable workflow for converting see-through XR hardware into a quantitatively specified ocular-light platform.

## 1 Introduction

Ocular light exposure is among the most powerful environmental regulators of human physiology [1, 2]. Beyond vision, light entrains the circadian clock [3, 4], suppresses pineal melatonin [5, 6], and modulates alertness [7]. Much of this non-visual response is mediated by melanopsin-expressing retinal ganglion cells (ipRGCs), which are intrinsically sensitive to light [1, 8, 9], and short-wavelength light around 460–480 nm is especially effective at suppressing melatonin [5, 6]. Describing such light therefore requires a standardized measure of melanopsin-effective light. The CIE S 026 *α*-opic system quantifies light according to the spectral sensitivity of the major retinal photoreceptors, and melanopic equivalent daylight illuminance (EDI) is its melanopsin-related quantity, a standardized measure of melanopsin-effective light at the eye. These measures describe the light stimulus reaching the eye rather than predicting any particular physiological response [1, 10], and consensus guidance recommends at least 250 lx melanopic EDI at the eye during the day and no more than 10 lx in the pre-sleep evening for healthy adults [2].

Translating this science into interventions is limited in part by exposure control. With light boxes, room lighting, or screens at a desk, the stimulus reaching the eye depends on viewing distance, gaze, posture, and ambient light, and individual sensitivity to evening light varies by more than tenfold [3, 11]. A light source worn on the head keeps the source at a more consistent position relative to the eyes, removing distance and posture as variables; gaze, pupil size, fit, ocular transmission, and ambient light can still change the light reaching the retina. This problem is especially relevant in light-poor clinical environments such as intensive care units (ICUs), where daytime illuminance in patient rooms is typically 74–160 lx [12–14] and melanopic EDI may remain well below daytime recommendations; in a newly built intermediate care unit, no 30-min average reached 250 lx [15]. Because physiological responses such as melatonin suppression are graded below those recommendations [11, 16], a worn source that adds a known, logged melanopic EDI spanning tens to a few hundred lux may still be experimentally and clinically relevant.

Head-mounted displays have recently been calibrated in detail and shown to deliver standardized light that reliably suppresses melatonin [17], but that work used fully immersive virtual-reality (VR) headsets that block the wearer’s view of the surrounding environment. Such occlusion is associated with cybersickness and related adverse effects [18] and prevents the wearer from seeing the surroundings, a drawback in supervised settings such as the ICU, where interest in controlled ocular light extends beyond circadian timing to physiological outcomes including autonomic and immune responses [19–21] and where light at the eye has been set by room geometry and patient position rather than controlled (Section 4.4). See-through extended-reality (XR) glasses overlay the display on a view of the real environment while keeping the source fixed relative to the head.

Consumer XR glasses are an attractive candidate: modern near-eye micro-OLED displays, one per eye, are bright, have separately addressable primaries, are light enough for chronic wear, and are inexpensive relative to laboratory photostimulators. They are, however, engineered as entertainment displays, not instruments. Operating-system color management, adaptive brightness, automatic brightness limiting (a reduction of display output under high load), undocumented optical geometry, and closed control stacks all stand between a requested pixel value and a known exposure at the eye, and none of them appears in a manufacturer’s specification, so the delivered stimulus has to be measured. For experimental use, however, characterization alone is insufficient: a usable platform must establish the mapping from software-controlled display settings to measured light exposure at the eye, quantify the prediction error of that mapping, and determine whether that calibration remains valid when software displays spatially structured application content.

Here we build and validate such a calibration pipeline, linking software control to measured ocular exposure, predictive modeling, and application-level estimates, for a representative commercial device, the VITURE Luma Ultra, in the sequence control, measure, characterize, calibrate, validate, and apply (Fig. 1). Relative to prior head-mounted work [17], the contribution is a see-through display with independently controlled and logged hardware intensity states, and a reusable calibration workflow consistent with calls for reproducible reporting of light interventions [22]. We combine controlled and logged display states with corneal-plane spectroradiometry, repeated device characterization, a settings-to-exposure model tested on held-out stimuli, and a test of how the calibration extends from uniform stimuli to rendered application content. The resulting workflow links software commands to measured ocular exposure, identifies which display settings and stimulus types support quantitative predictions, and can be applied to other head-worn displays.

**Figure 1:**
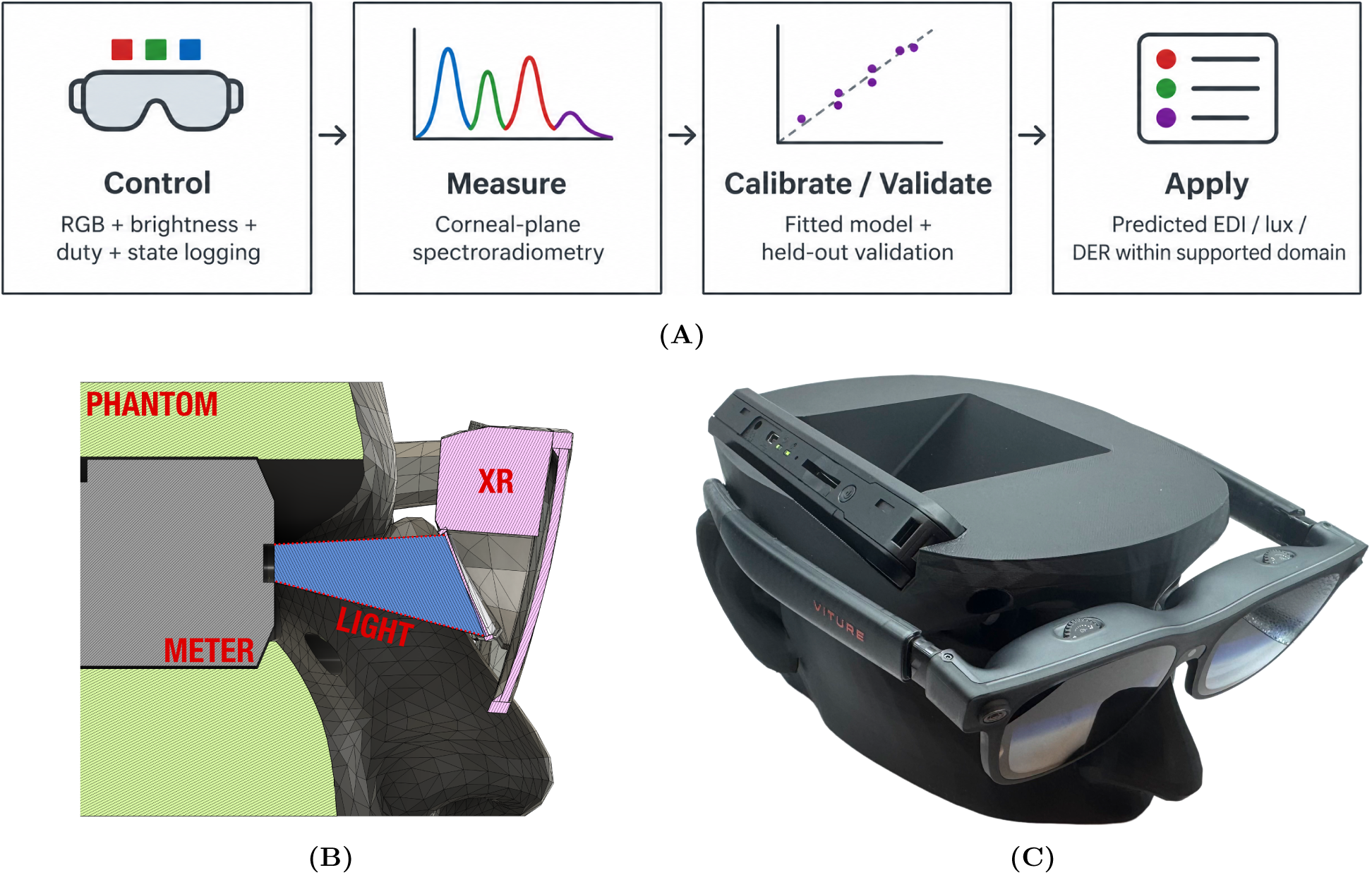
Calibration pipeline and measurement geometry. (**A**) Workflow from controlled display state to corneal-plane measurement, calibration and validation, and application prediction. (**B**) Cross section of the phantom-head measurement fixture (Fusion 360), with elements labeled in red: PHANTOM, the phantom head; METER, the spectroradiometer, with its diffuser at the corneal plane; XR, the glasses’ display module for the right eye; and LIGHT, the light cone from the display to the diffuser, whose dotted margins mark the shortest and longest geometric distances (Table 1). (**C**) Printed phantom (black PLA) with the spectroradiometer installed and the VITURE Luma Ultra glasses in the worn position.

**Table 1:** Measurement instrument and configuration. Specifications are from the manufacturer [25].

| Element | Specification |
| --- | --- |
| Spectroradiometer | UPRtek MK350S Premium (reports as MK350X; firmware 1.1.1.B4) |
| Spectral range | 380–780 nm, 1 nm data increment; bandwidth approximately 9 nm half bandwidth |
| Wavelength reproducibility | $\pm 1$ nm |
| Measurement range | 1 to 150,000 lx; integration time 60 μs to 5,000 ms |
| Illuminance accuracy | $\pm 2.5\%$ (CIE Illuminant A, 2,856 K) |
| Stray light | –25 dB max. |
| Cosine response | JIS C 1609-1:2006 General Class AA; DIN 5032-7 Class B |
| Quantity | Cosine-corrected spectral irradiance at the right eye position (both displays lit) |
| Probe placement | stock MK350S Premium diffuser at the corneal plane of the right eye; geometric distance from the right display 31.19–39.68 mm (mean 35.44 mm; Fig. 1B); meter held in a custom cutout of the phantom head with the diffuser at the corneal plane |
| Diffuser | active diameter 7 mm; centered on the gaze axis; normal to the gaze axis, facing the display |
| Exposure | auto-exposure, ceiling 500 ms; dark calibration with capped sensor at the start of each session |
| Capture | 3 sessions; 3 repeats per stimulus; settle 1 s (3 s after a brightness change) |

## 2 Materials and Methods

### 2.1 Display platform and stimulus host application

The stimulus platform is a pair of VITURE Luma Ultra extended-reality glasses: a binocular micro-OLED near-eye display (1920*×*1200 pixels per eye, nine discrete hardware brightness levels indexed 0–8, and an electrochromic dimming film) [23]. It was driven over USB-C from a Microsoft Windows 11 host as an extended display in the two-dimensional 1920*×*1080 at 60 Hz mode, in which both eyes are shown the same frame (glasses firmware 12.0.01.101_20260605; VITURE Glasses SDK 2.4.0). How that frame is mapped onto the 1920*×*1200 panels was not determined; because the mapping can change the emitting area, the measurements characterize the tested display mode.

Stimuli are presented by a purpose-built host application (chronolume_host) that renders the requested 8-bit RGB pixel values (“codes”; 0–255 per channel) in a full-screen window on the glasses, with HDR, Night light, and automatic color management turned off for the glasses display. Before every capture, the application reads the rendered framebuffer back from memory to verify that the requested pixel values were actually rendered (Section 2.3). Framebuffer readback verifies the application framebuffer but cannot exclude subsequent processing by the operating system, display interface, or glasses electronics, so the emitted output was characterized optically. A separate USB interface through the manufacturer’s SDK library sets and reads hardware state that is independent of pixel values: the brightness level (0–8), the electrochromic film state, the panel duty cycle, and the display mode [24]. The duty cycle, which the documentation describes as the percentage of time the display is on, is limited to the manufacturer’s presets (30, 42, and 98); its optical effect was checked qualitatively with a high-speed optical sensor (Section 2.3). Each state is read back after it is set. The duty cycle was held at 98 except during the duty series, and the film was off throughout. Every stimulus change is logged with the requested RGB code, brightness level, duty cycle, film state, display mode, and calibration identifier, so that the hardware states relevant to intensity are recorded with the stimulus and frozen during a sweep. Host hardware, renderer settings, and the remaining configuration are listed in Appendix C.

### 2.2 Optical measurement

Spectral irradiance was measured with a handheld spectroradiometer (UPRtek MK350S Premium; Table 1) [25], which reports factory-calibrated, cosine-corrected spectral irradiance from 380 to 780 nm in 1 nm increments. The meter was held in a phantom head (Fig. 1B,C) with a custom cutout that places its stock diffuser, 7 mm in active diameter, at the corneal plane of the right eye, centered on the gaze axis and oriented normal to the gaze axis, facing the display, with the glasses in the worn position on the phantom (geometry in Table 1). The measured quantity is therefore corneal-plane spectral irradiance *E_λ_*at the right eye with both displays lit. This is the quantity in which CIE S 026 and the consensus recommendations are written [2, 10], so no field-of-view or geometric assumption enters any reported value.

Measurements were made in a dark room, with a black curtain over the phantom during measurements. The black stimulus (0, 0, 0) read 0.033–0.087 lx across sessions (mean 0.059 lx, 0.06 lx melanopic EDI), which combines residual room light, the panel black level, and the meter’s dark noise. The black stimulus therefore captures residual room light, display black-level emission, and instrument dark noise, and characterization values are reported without subtracting this black measurement. Readings below the meter’s specified photopic minimum of 1 lx [25] are reported but should be interpreted as approximate.

### 2.3 Automated acquisition and quality control

A measurement package drives the host application and captures with the meter through the vendor’s USB library. After a 10-min warm-up of the glasses, three sessions were run in succession (5.8–6.6 min each, 22 min in total). Before each session the glasses were removed and reseated on the phantom, and each session began with its own dark calibration with the sensor capped. Each measurement capture consisted of 3 repeated meter readings at one display setting, and each session comprised the same 62 captures, characterizing the primary and white spectra, a six-point blue code series, intermediate codes for the other channels, all hardware brightness levels, and all available duty-cycle settings; the complete stimulus sequence is given in Appendix C. Brightness and duty cycle were changed only while the display was black and were read back before each capture; steps without their own brightness level or duty cycle ran at level 8 and duty 98. Full white at brightness level 8 produced a reproducible display and control failure on more than one host and was treated as an unsupported operating point, so white measurements were limited to level 7 (evidence in Appendix C). The meter used auto-exposure with a ceiling of 500 ms (exposures ranged from 6 to 500 ms), and the dark calibration ran under the same ceiling. At that ceiling, (0, 0, 16) and black (0.13 and 0.059 lx photopic) did not reach the meter’s minimum signal and were flagged out of range in every session; they are reported as readings below the meter’s range, like every value below 1 lx. Each capture waited 1 s after the stimulus change (3 s after a brightness change).

Quality control uses only each capture’s own readings, never a comparison with an expected value: every capture required a framebuffer readback that matched the requested code, detectable display light, stable repeats, a stable repeated reference stimulus (“anchor”; full-code blue at brightness 8, captured at the start and after every five steps), and agreement between the photopic illuminance computed from the exported spectrum and the meter’s own; failed checks were retaken or their measurement block (the stimuli at one brightness level and duty cycle, bracketed by black captures) re-run (criteria and retry rules in Appendix C). Sessions containing unresolved QC failures were excluded from quantitative analysis. All raw readings, discarded readings, and host state are stored with each capture.

#### Optical duty cycle

Because the meter integrates over each exposure, the temporal meaning of the duty-cycle setting was checked separately with a high-speed optical sensor (OPT101, Texas Instruments) recorded on a logic analyzer (Logic Pro 8, Saleae) across duty cycles, colors, brightness levels, and the 60 Hz and 120 Hz display refresh settings. This check was qualitative; the recordings are not reported as data.

Each reported value is the mean over the three sessions, with the between-session range where shown. Because the sessions differed in dark calibration and in the seating of the glasses on the phantom, as well as in time, they are treated as repeated measurements and no trend over time is estimated. Repeatability is summarized at three scales: within a capture, by the coefficient of variation (CV) of integrated irradiance across its repeats; within a session, by the anchors; and between sessions, by the CV of each capture’s photopic illuminance across sessions.

### 2.4 Photometric, *α*-opic, and safety quantities

All quantities are corneal-plane exposure intensities (irradiance, illuminance, EDI); a time-integrated dose would additionally require the exposure duration. Photopic illuminance and the *α*-opic quantities, including melanopic EDI and the melanopic daylight efficacy ratio (DER, melanopic EDI divided by photopic illuminance), were calculated according to CIE S 026 [10]. Appendix C gives the definitions and the spectral summary metrics (integrated, band, and photon irradiance, peak wavelength, FWHM, centroid, and band shares).

The response to blue code was summarized by a power law, *E*(*B*) = *E*_255_ (*B/*255)*^γ^*, fitted in log–log space to the 380–500 nm band irradiance of the six blue codes in each session. Both *E*_255_ and *γ* are fitted parameters: *E*_255_ is the fit’s value at code 255, not the measured code-255 value, so the measured value can deviate from it. Because no change in spectral shape with code was resolved (Section 3.2), the same exponent applies to melanopic EDI. For comparison across channels, the same form was fitted to the session-mean photopic illuminance of codes 64–255 of each channel. The response to hardware brightness was not fitted with a function: each level is reported as measured, with the ratio of its photopic illuminance to that of the level below. The response to duty cycle is reported as each channel’s melanopic EDI relative to its value at duty 98, with the blue peak wavelength and FWHM at each duty.

Photobiological safety was screened from the blue-light-hazard weighted irradiance *E_B_* = *E_λ_B*(*λ*) *dλ*, using the ICNIRP *B*(*λ*) weighting, against the ICNIRP small-source blue-light-hazard criterion *E_B_ ≤* 1 W m^−2^ for exposures longer than 100 s [26, 27]. Because the measured irradiance integrates the whole display over a diffuser whose active diameter (7 mm) equals the ICNIRP averaging aperture, the comparison is conservative with respect to the angular distribution of radiance and needs no assumption about the homogeneity of the irradiance profile (Appendix C).

### 2.5 Settings-to-exposure model

The characterization above measures a fixed set of stimuli. To let software compute the exposure of any uniform stimulus from its settings alone, a forward prediction model was calibrated to map display settings to corneal melanopic EDI and photopic illuminance, and from them the melanopic DER, for any code (*R, G, B*), hardware brightness level *L*, and duty cycle *D* within the calibrated range.

#### Acquisition

A dedicated sweep used the same rig, host application, and quality control as the characterization, with uniform full-frame stimuli in four calibration stages: primary calibration, each primary alone; RGB code series measured at multiple brightness levels, added because the code response changed with level; mixture calibration, 19 two- and three-channel compositions including the gray axis and full white, at level 7 or below; and a supplemental mixture-calibration stage acquired before validation. Each stage was repeated in three sessions (the supplemental mixture stage in two) with the glasses reseated between sessions; the full-code output at the remaining brightness levels comes from the characterization sessions, and two code-series sessions in which the glasses had slipped on the phantom were excluded before fitting. A separate validation session, not used for fitting or model selection, contained previously unmeasured codes, levels, and compositions that are not in the calibration grid. Duty cycle could not be held out, because the device offers only three duty-cycle presets. Settings were grouped into measurement blocks of one level and duty cycle, each bracketed by black captures, in randomized order, so the hardware state never changed inside a block. The complete grids are given in Appendix Table 5 and the design rationale in Appendix A.1.

#### Model

The display-attributable output of each setting, defined as its measured value after subtracting the mean of the two black captures bracketing its block, is the quantity modeled. For each primary *c* and output, a separable (factorized) model, in which code, level, and duty cycle each scale the output independently, takes the form

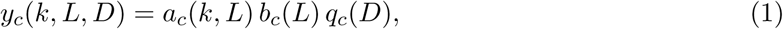

a code response *a_c_* (1 at code 255), a full-code level response *b_c_*, and a duty response *q_c_*(1 at the highest calibrated duty), fitted by least squares in log space. The code response depends on level, so it is fitted at each of several levels of the code series and interpolated between them. The model does not extrapolate outside its calibrated domain in level, duty cycle, or mixture level, and returns no value there; the one exception is in code, below a level’s lowest code node, where the lowest segment is extended toward code 0 and the prediction is flagged as extrapolated. A stimulus is predicted as the sum of its primaries. Because mixtures were not additive (in the calibration mixtures a dim channel next to a full-code channel lost output), candidate models could include a mixture-interaction term that reduces a primary’s predicted contribution in the presence of other channels. The DER is the ratio of predicted melanopic EDI to predicted photopic illuminance. Appendix A.1 gives the code nodes, the interaction terms, the crossed alternative form, and the rationale for each term.

#### Selection and evaluation

Seven candidate model structures differing in factorization, mixture interaction, and treatment of low-signal red melanopic output were compared by leave-one-session-out cross-validation on the calibration sessions, refitting every parameter in each fold (Appendix A.1). The candidate with the fewest failed predictions, and among those the lowest worst-case cross-validation error, was selected; the selected model and its parameters were fixed before the validation session was examined. Prediction error is reported as the mean absolute percentage error (MAPE), the 95th-percentile absolute percentage error (P95), and the mean signed percentage error (bias), for melanopic EDI and photopic illuminance separately. No prespecified validation-performance criterion was used to declare the model successful; the fitting software’s model-development preference thresholds, which no candidate met, are described in Appendix A.1. Eligibility was decided by the measurement alone, separately for each output: a setting was eligible if its photopic illuminance before black subtraction (“gross” illuminance) was at least 1 lx, the meter’s specified minimum, and its display-attributable output exceeded 3 times the variation of its bracketing black captures (Appendix A.1). A failure is an eligible setting for which the model returns no value (outside its supported domain); failures are counted separately, and percentage errors are computed over the successful predictions. The predicted value never decides eligibility. The locked model and its configuration (device, firmware, film state, display mode, and calibration identifier) are exported as a versioned calibration package. Appendix A gives the complete prediction algorithm, the fitted parameters, and a worked example.

### 2.6 Example application frames

The calibration covers uniform full-frame colors, but an application shows composed frames. To test whether the uniform-field calibration could be extended to spatially structured application content, we asked how far a frame’s exposure can be estimated from its pixels. Four example frames, following a mockup of a photostimulation application, were rendered at the framebuffer size (Fig. 5A; exact colors and pixel fractions in Appendix C): a morning text slide and a blue orb at level 8, and a night text slide and a red orb at level 5. The orbs and the morning slide were composed to the calibrated domain, with pure primaries at calibrated codes on a black background. The night slide was taken as designed, so its background, 88% of the frame, lies below the lowest calibrated code (64 for red, green, and blue) and its prediction is extrapolated.

Each frame was predicted without using the frame sessions, at its level and duty cycle 98: the locked model of Section 2.5, with the code response at the frame’s level and the mixture interaction, was evaluated for each of the frame’s colors and summed over its color histogram (the fraction of pixels at each RGB value), weighted by pixel fraction (equal spatial weighting; extrapolation as in Step 2 of Appendix A). For comparison, each frame was also predicted as a uniform field of its mean RGB code, a natural shortcut for a developer, which the nonlinear code response makes biased.

The frames were shown pixel for pixel by the host application, which reads back the whole framebuffer and compares it with the frame; a frame was measured only if every pixel matched. Each frame was measured in 3 sessions, with the glasses reseated between them, together with each of its non-black colors as a uniform full-field reference (block order in Appendix C); frame and references are net of the block’s black captures. The pixel-weighted sum of the same-session references tests spatial additivity, that is, whether a rendered frame produces the pixel-fraction-weighted sum of the corresponding uniform-field measurements, without any model: a frame that equals it behaves as the sum of its uniform parts.

#### Spatial grid

The frames turned out not to be additive in this sense (Section 3.5), so a spatial grid was measured afterwards to find out why. The frame was divided into a 3 *×* 3 grid of equal cells; each cell was lit alone at full code, and so was the full field, for red, green, blue, and white at level 7 and duty cycle 98, in 3 sessions with the glasses reseated between them, using the same block structure, black subtraction, and anchor as the frames. Nine times a cell response divided by the full-field response measures the combined effect of spatial weighting and the lit-area gain, which we define as the increase in per-pixel output when only a fraction of the display is illuminated. The relative pattern across the cells identifies spatial weighting (uniform for equal weighting); the sum of all nine cells relative to the full field quantifies the lit-area gain (1 for an additive display whatever the weighting; above 1, a small lit area gives more light per pixel than a full field).

#### Spatial correction model

From the grid alone, without the frame sessions, a spatial weight map and a lit-area gain were derived and applied to each color’s uniform prediction, and to the same-session references (Appendix A.1). This secondary spatial correction accounts for position-dependent weighting and lit-area gain. It was formulated after the frame measurements but derived entirely from the independent spatial-grid data, with none of its parameters fitted to the frame measurements; it was therefore assessed as a consistency test rather than an independently validated model. It describes the phantom and probe rather than a wearer’s eye, and is reported alongside the equal-weighting estimate, outside the validated uniform-field model.

## 3 Results

### 3.1 Display control and measurement repeatability

Application framebuffer readback of both halves matched the requested code for all 228 captures across the three sessions (186 stimuli and 42 anchors), with no corrective action needed. The brightness level and duty cycle read back from the glasses matched the requested values, and the display mode and film state were constant throughout. No reading was discarded for lack of display light, and no capture or block needed to be re-run. Readback establishes the codes in the application framebuffer, not the values reaching the panel; the optical measurements show that the emitted output rises with the requested code and hardware brightness level (levels 0 and 1 were indistinguishable within repeatability) with no resolved change in spectral shape (Sections 3.2 and 3.3).

Within a capture, repeats agreed to 0.3% CV or better for every stimulus within the meter’s measurement range. Within a session, the 14 anchors of each session stayed within 4.0% of that session’s first anchor. Between sessions, which differed in dark calibration and in the seating of the glasses as well as in time, the photopic illuminance of each in-range capture varied with a median CV of 1.9% and at most 2.6%, and the first anchors of the three sessions spanned 3.5% of their mean. The melanopic EDI of full blue at level 8 varied by 2.2% CV. The between-session spread is about an order of magnitude larger than the within-capture spread and is the largest observed component of measurement variability. It combines differences in seating, dark calibration, and display output, which were not separated. These figures describe repeatability, not absolute accuracy, which is bounded by the meter and the geometry (Section 4.5).

### 3.2 Spectral characterization

The native blue primary peaks at 459 nm with a measured FWHM of 34.7 nm and a centroid of 469 nm (Table 2; Fig. 2). The measured width includes the meter’s spectral bandwidth (approximately 9 nm half bandwidth in the archived datasheet and 12 nm on the current product page [25]), so the intrinsic emission width is somewhat narrower; we report the measured value. Of the 380–780 nm irradiance, 95.5% falls within 380–500 nm; the remaining 4.5% is split between 500–600 nm (2.8%) and 600–780 nm (1.7%).

**Table 2:** Spectral characterization of the display primaries at their highest usable brightness level (corneal-plane irradiance at the right eye position; means of three sessions). The FWHM of white is not given because its spectrum has several peaks.

| Stimulus | Level | Peak<br>(nm) | FWHM<br>(nm) | Centroid<br>(nm) | Band share (%) |  |  |
| --- | --- | --- | --- | --- | --- | --- | --- |
|  |  |  |  |  | 380–500 | 500–600 | 600–780 |
| R (255,0,0) | 8 | 631 | 78 | 642 | 0.1 | 13.4 | 86.5 |
| G (0,255,0) | 8 | 539 | 86 | 549 | 10.6 | 79.3 | 10.2 |
| B (0,0,255) | 8 | 459 | 35 | 469 | 95.5 | 2.8 | 1.7 |
| W (255,255,255) | 7 | 459 | – | 560 | 27.2 | 40.4 | 32.4 |

**Figure 2:**
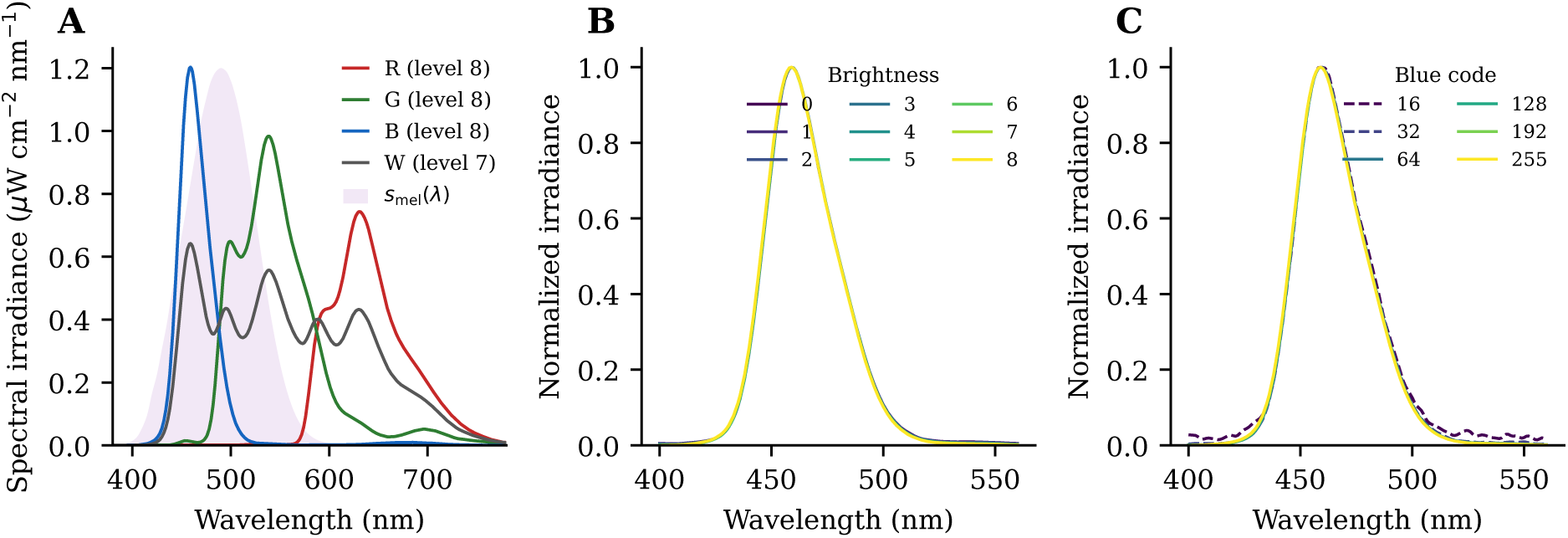
Display spectra at the corneal plane (means of three sessions). (**A**) Spectral irradiance of full-code red, green, and blue at brightness level 8 and white at level 7, with the CIE S 026 melanopic action spectrum (scaled) for reference. (**B**) Full-code blue at brightness levels 0–8, normalized to peak. (**C**) Blue codes 16–255 at brightness 8, normalized to peak; codes 16 and 32 (dashed) are below the meter’s specified 1 lx photopic minimum and are approximate.

No change in spectral shape was detectable across intensity, within the measurement resolution and repeatability. Across hardware brightness levels 0–8 and blue codes 32–255, in every session, the peak stays at 459–459 nm and the FWHM at 34.4–34.8 nm (Fig. 2B,C). Code 16 is excluded from this range because its reading falls below the meter’s specified range (Section 3.3). Intensity can therefore be manipulated without altering the spectral stimulus. The peak lies within the short-wavelength band relevant to melanopsin (the CIE S 026 melanopic action spectrum peaks near 490 nm [10]).

The three primaries have distinct spectral peaks (Fig. 2A; Table 2): red peaks at 631 nm and green at 539 nm, with 86.5% and 79.3% of their irradiance in their own bands. White combines all three and spans the visible range. The green primary is broad (FWHM 86 nm) and produces substantial melanopic stimulation: its melanopic DER is only 0.63, but because it is much brighter photopically it reaches 248 lx melanopic EDI at level 8, matching the blue primary’s 252 lx at the same level. White is not the sum of its primaries. Measured at the same brightness level in the brightness series, full white reached 89% of the summed photopic illuminance of red, green, and blue at level 0 and 81% at level 7, with the shortfall growing with output. This is consistent with panel-level power limiting, although the mechanism was not examined, and with the failure of white at level 8. Per-channel calibrations therefore cannot be added linearly to predict mixed stimuli; mixtures must be measured directly or predicted by a model calibrated on measured mixtures (Section 3.4).

### 3.3 Intensity control and usable exposure range

The three intensity controls behaved differently and were therefore characterized separately, and all are logged with every stimulus. Along the digital-code control axis (the code axis), melanopic EDI of the blue primary rises from 0.71 lx at code 16 to 252 lx at code 255 (Fig. 3A). A power law summarizes it with *γ* = 2.04 in every session (*R*^2^ *≥* 0.9990 in log space), but the deviations from the power-law fit followed the same pattern in all three sessions: the mid codes lie above the fit (code 64 by 5%) and the ends below it (code 255 by -5%), with a largest mean deviation of 5%. The curvature is therefore a property of the display rather than noise, and the code axis, like the brightness axis, is best used as a measured lookup. Red, green, and blue follow similar curves: fitted to codes 64–255, their photopic illuminance rises with exponents of 1.94–1.99, and white with 2.22. The low end is limited by the measurement rather than the display: blue codes 16 and 32 fall below the meter’s specified 1 lx minimum, so values for those codes should be interpreted as approximate.

**Figure 3:**
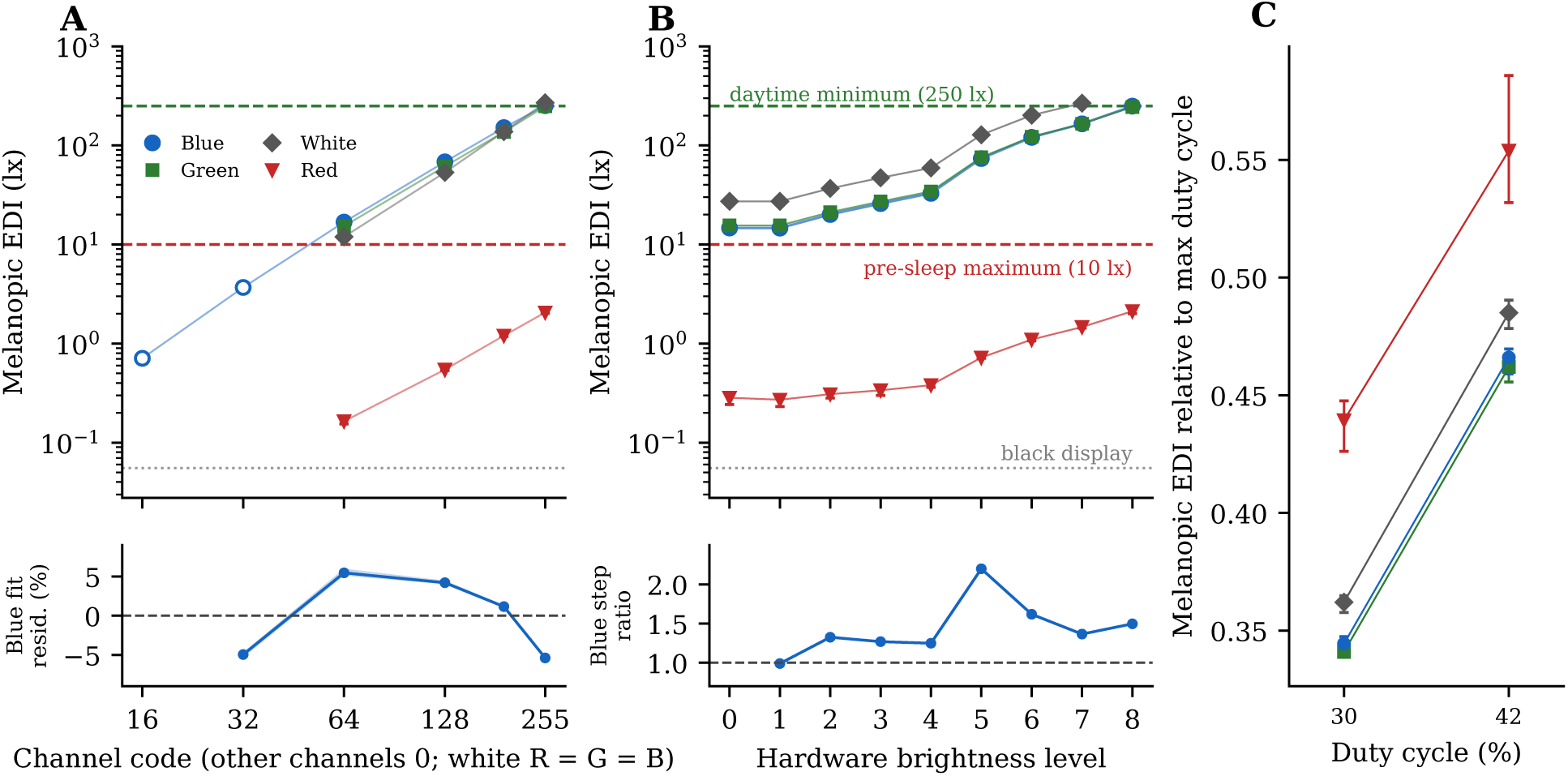
Corneal melanopic EDI along the three intensity axes, all channels. Markers are means of three sessions; error bars give the between-session range and are mostly smaller than the markers. (**A**) Melanopic EDI versus code of each channel (other channels 0; white has equal codes); red, green, and blue are at brightness 8 and white at 7. The strip below gives the mean (line) and between-session range (band) of the percent deviation of the blue points from each session’s own power-law fit (*E*_255_(*B/*255)*^γ^*, *γ* = 2.04, codes 32–255), showing the same curvature in every session. Open markers (blue codes 16 and 32) are readings below the meter’s specified 1 lx photopic minimum and are approximate. (**B**) Melanopic EDI of each channel at full code across hardware brightness levels (white to level 7); the strip below gives the mean (line) and between-session range (band) of the ratio of full-code blue photopic illuminance at each level to that at the level below. In A and B, dashed lines mark the consensus daytime minimum (250 lx) and pre-sleep maximum (10 lx) for healthy adults [2], and the dotted line the black display. (**C**) Melanopic EDI of each channel at full code (red, green, and blue at level 8, white at level 7) at the lower preset duty cycles (42 and 30), relative to its value at the maximum duty cycle of 98 (1 by definition, not shown). Colors and markers in all panels are as in the legend of A.

Along the hardware-brightness control axis (the brightness axis), melanopic EDI of full-code blue in the brightness series rises from 14.7 lx at level 0 to 249 lx at level 8, a 17.0-fold range (Fig. 3B; Table 11). Output rises with level but not uniformly: levels 0 and 1 were indistinguishable within repeatability (illuminance step ratio 0.990), and the largest step is from level 4 to 5 (*×*2.20). No smooth function describes it well, so the level axis is used as a measured lookup table. The normalized brightness response is similar across primaries: at every level, the illuminance of red, green, and blue as a fraction of its own level-8 value agreed within 7%, and the 4-to-5 step was *×*2.12–2.20 for all four channels. The lookup values themselves are channel-specific (Table 11). White follows the same steps at low levels but showed progressively less output than expected from the individual primaries at the highest brightness levels, as expected from its growing shortfall from additivity. Full-code blue at level 8 was captured twice per session, once in the primary and code series (Table 3) and once in the brightness series (Table 11), and the session means agreed within 1.1%. Duty cycle, the third intensity control, follows.

**Table 3:** Corneal-plane exposure intensity at the right eye position, measured (Level: hardware brightness; duty cycle 98). Values are means of three sessions; the range is the between-session range of melanopic EDI. Red, green, and blue are at level 8 and white at level 7, the highest usable level for each, from the primary capture series (Section 2.3).

| Stimulus | Level | Illuminance<br>(lx) | Irradiance ($\mu\text{W cm}^{-2}$) | | Photon irradiance ($10^{14} \text{ cm}^{-2} \text{ s}^{-1}$) | | Melanopic EDI (lx) | | S-cone-opic<br>EDI (lx) | Melanopic<br>DER |
| --- | --- | --- | --- | --- | --- | --- | --- | --- | --- | --- |
|  |  |  | 380–780 | 400–500 | 380–780 | 400–500 | mean | range |  |  |
| R (255,0,0) | 8 | 115 | 61.4 | 0.0618 | 1.99 | 0.00146 | 2.06 | 2.02–2.09 | 0.453 | 0.018 |
| G (0,255,0) | 8 | 392 | 81.4 | 9.27 | 2.25 | 0.229 | 248 | 243–254 | 30.9 | 0.63 |
| B (0,0,255) | 8 | 30.0 | 45.9 | 44.0 | 1.08 | 1.03 | 252 | 246–257 | 376 | 8.38 |
| W (255,255,255) | 7 | 296 | 102 | 28.1 | 2.87 | 0.663 | 269 | 263–275 | 214 | 0.91 |

Lowering the duty cycle from 98 to 42 and 30 reduced output without changing the spectrum, but output decreased less than would be expected if optical output scaled directly with the duty-cycle setting (Fig. 3C; Table 12): the melanopic EDI of every channel was above proportional scaling, by up to 43.6% (for red). For blue at duty 30 it fell to 0.344 of its value at duty 98, against 0.306 for proportional scaling, while the blue peak moved by at most 0 nm and its FWHM by at most 0.1 nm. Duty is therefore a third logged intensity axis, independent of code and hardware brightness, whose effect on exposure must be taken from the measured values rather than from the setting.

The optical duty cycle was confirmed qualitatively to track the requested duty setting with the OPT101 sensor (Section 2.3): the emitted waveform changed with the setting at both the 60 Hz and 120 Hz refresh settings, with a period consistent with the refresh rate in use, and it was not visibly affected by RGB code or brightness level. The read-back value confirms only that the SDK accepted the setting. Because this check was qualitative, the effect of duty on exposure is taken from the measured values above.

Table 3 reports the measured corneal-plane quantities. Full-code blue at maximum brightness gives 30.0 lx photopic but 252 lx melanopic EDI (246–257 lx across sessions; melanopic DER 8.38), with a corneal irradiance of 45.9 *µ*W cm^−2^ over 380–780 nm, of which 44.0 *µ*W cm^−2^ falls in 400– 500 nm (photon irradiance in Table 3; further spectral quantities in Table 2). Full green at level 8 gives a similar 248 lx (243–254 lx) at more than ten times the photopic illuminance (392 lx). Full white at level 7 gives the highest melanopic EDI, 269 lx (263–275 lx), at 296 lx photopic.

Against the consensus recommendations [2], full white at its highest usable level reached the 250 lx daytime minimum in 3 of 3 sessions, and full blue at maximum reaches 101% of it. At the other end, even the dimmest brightness level of full-code blue (14.7 lx) exceeds the 10 lx pre-sleep maximum, so evening-compatible blue exposures require lowering the code as well (code 64 at level 8 gives 17 lx and code 32 approximately 3.7 lx), or a shorter duty cycle: duty cycle 30 scales blue to 0.33 of its output at duty 98, so that full-code blue at level 4 and duty 30, a calibration setting (Section 2.5), is predicted at 10.7 lx, near the pre-sleep maximum. Combinations of code, level, and duty cycle between the calibrated ones are predicted by the exposure model (Section 3.4). The corneal blue irradiance is above the monochromatic short-wavelength exposures (roughly 1– 30 *µ*W cm^−2^ at 446–477 nm) that produced melatonin suppression, circadian phase shifts, and alerting effects in controlled laboratory studies [5–7], although those exposures filled a much larger part of the visual field.

The blue-light-hazard weighted irradiance of full-code blue at brightness 8 is 0.31 W m^−2^, a factor of 3 below the small-source criterion of 1 W m^−2^ [26, 27]; full white at level 7 gives 0.18 W m^−2^ (a factor of 5 below) and full green at level 8 0.037 W m^−2^ (a factor of 27 below). These values are averaged over the 7 mm ICNIRP aperture (Section 2.4). Under this conservative screening the tested stimuli pass; a full extended-source assessment would require the angular radiance distribution of the display, which was not measured.

### 3.4 Prediction of exposure from display settings

Cross-validation on the calibration sessions selected the factorized model with a per-channel mixture-interaction term (Appendix A.1). Because red melanopic output was near the measurement floor at low intensities, red melanopic EDI was estimated from its photopic prediction and a calibrated melanopic DER. The model structure and parameters were fixed before the independent validation session was examined. On the 48 validation settings (codes, levels, and mixtures not in the calibration grid, at calibrated duty cycles; Table 4; Fig. 4) it predicted melanopic EDI with a MAPE of 3.4% (95th percentile 8.8%, bias 0.3%, mean absolute error 0.39 lx), photopic illuminance with a MAPE of 4.5% (95th percentile 10.3%, bias 3.5%, mean absolute error 0.38 lx), and the melanopic DER with a MAPE of 3.5%, with 0 failed predictions among the 34 settings eligible for melanopic EDI and 0 among the 41 eligible for photopic illuminance. For the mixtures alone the MAPE was 2.9% for melanopic EDI and 4.3% for photopic illuminance (by group in Table 7 and by composition in Table 8, Appendix A). Prediction errors showed little systematic dependence on brightness level or duty cycle: at every validation level and duty cycle the mean signed error of each output was within 5% (Fig. 7, Appendix A). The photopic bias was concentrated in the 27 settings with a channel at code 96 (mean signed error +5%, against +1% for the others), a code interpolated between the calibrated code nodes 64 and 128. The largest error was +12% for (96, 0, 0) at level 6 and duty 42. For context, the meter’s specified illuminance accuracy is *±*2.5% (Table 1), and the median between-session coefficient of variation of the characterization sessions was 1.9%.

**Table 4:** Prediction error of the settings-to-exposure model on the independent validation session (codes, levels, and mixtures not in the calibration grid, at calibrated duty cycles) and in leave-one-session-out cross-validation on the calibration sessions, for all stimuli (by group in Table 7). MAPE: mean absolute percentage error; P95: 95th percentile of absolute percentage error; bias: mean signed percentage error (all over the *n* successful predictions); MAE: mean absolute error in lx over every quantitative setting for which the model returned that output, eligible or not (not applicable to the dimensionless DER). *n* counts the settings eligible for that output (for the DER, eligible for both, decided by the measurement alone; Section 2.5) that received a prediction. Not eligible: 14 settings for melanopic EDI and 7 for photopic illuminance. Failed (eligible, no prediction): 0 for melanopic EDI and 0 for photopic illuminance. Eligible settings with predicted photopic illuminance below 1 lx, included above: 1.

| Output | Independent validation |  |  |  |  | Cross-validation |
| --- | --- | --- | --- | --- | --- | --- |
| | $n$ | MAPE (%) | P95 (%) | Bias (%) | MAE (lx) | MAPE (%) |
| Melanopic EDI | 34 | 3.4 | 8.8 | 0.3 | 0.39 | 6.1 |
| Photopic | 41 | 4.5 | 10.3 | 3.5 | 0.38 | 4.9 |
| Melanopic DER | 34 | 3.5 | 8.6 | -2.4 | – | 4.5 |

**Figure 4:**
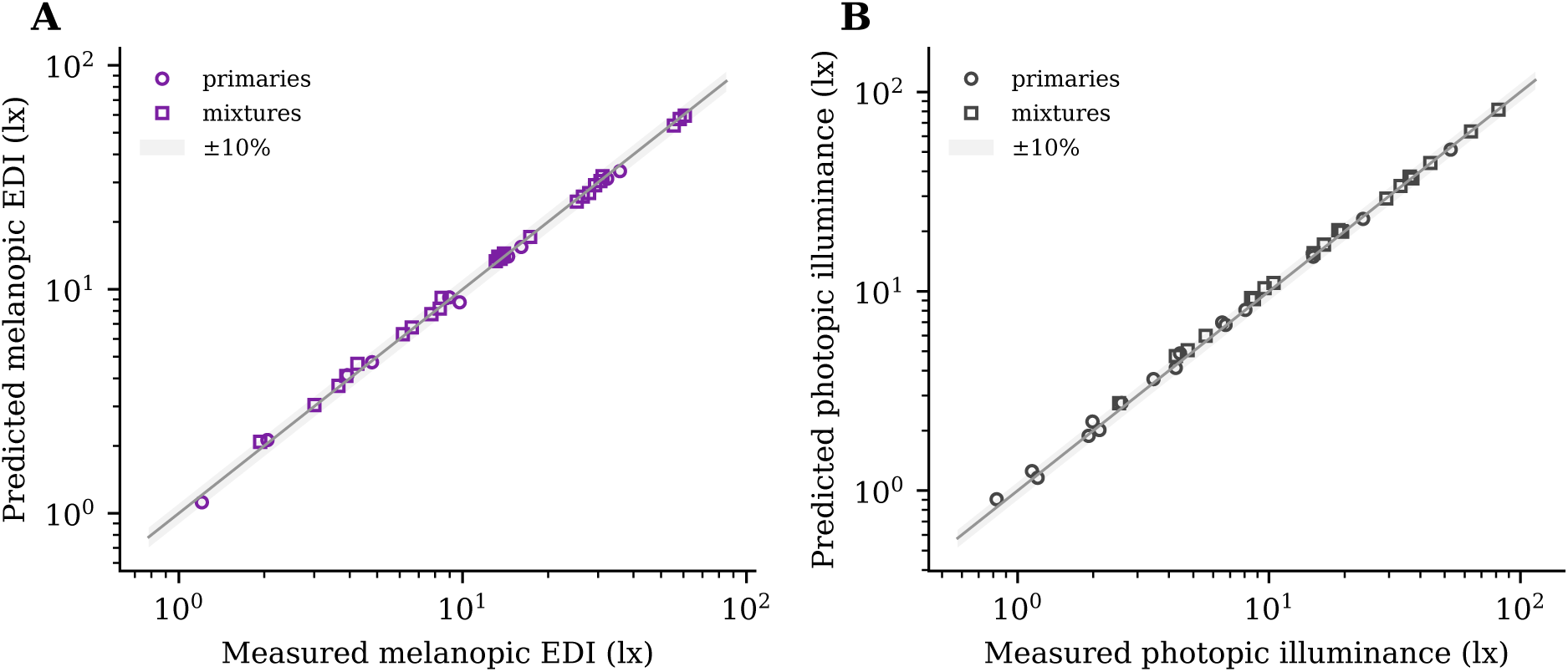
Independent validation of the settings-to-exposure model. Predicted versus measured (**A**) melanopic EDI and (**B**) photopic illuminance on the held-out validation session, for single-primary (circles) and mixed (squares) stimuli eligible for that output; the line is identity and the band *±*10%. Error statistics are in Table 4.

### 3.5 From uniform stimuli to rendered application frames

The frames test where the uniform-field calibration stops applying (Fig. 5; absolute values in Table 13). Predicting a frame as a uniform field of its mean RGB code failed outright, because the code response is strongly nonlinear: melanopic EDI was underestimated 2.7-fold for the blue orb and 3.9-fold for the red. Thus, at minimum, predicting rendered content requires the distribution of RGB values across the frame rather than its mean RGB value. Predicted per pixel from the histogram with the locked model, the frames were still low by about a third in melanopic EDI (Fig. 5C): by -35%, -35%, -24%, and -36% for the morning slide, blue orb, night slide, and red orb. This discrepancy was not primarily attributable to model error: the pixel-weighted sums of the same-session measured uniform references, which involve no model, were low by about as much (−37%, -36%, -35%, and -34%), so the frames are not additive in their uniform parts; each frame gave 1.44–1.60 times the photopic illuminance of its reference sum. The night slide’s extrapolated background (88% of the frame) contributed 2% of its predicted melanopic EDI.

**Figure 5:**
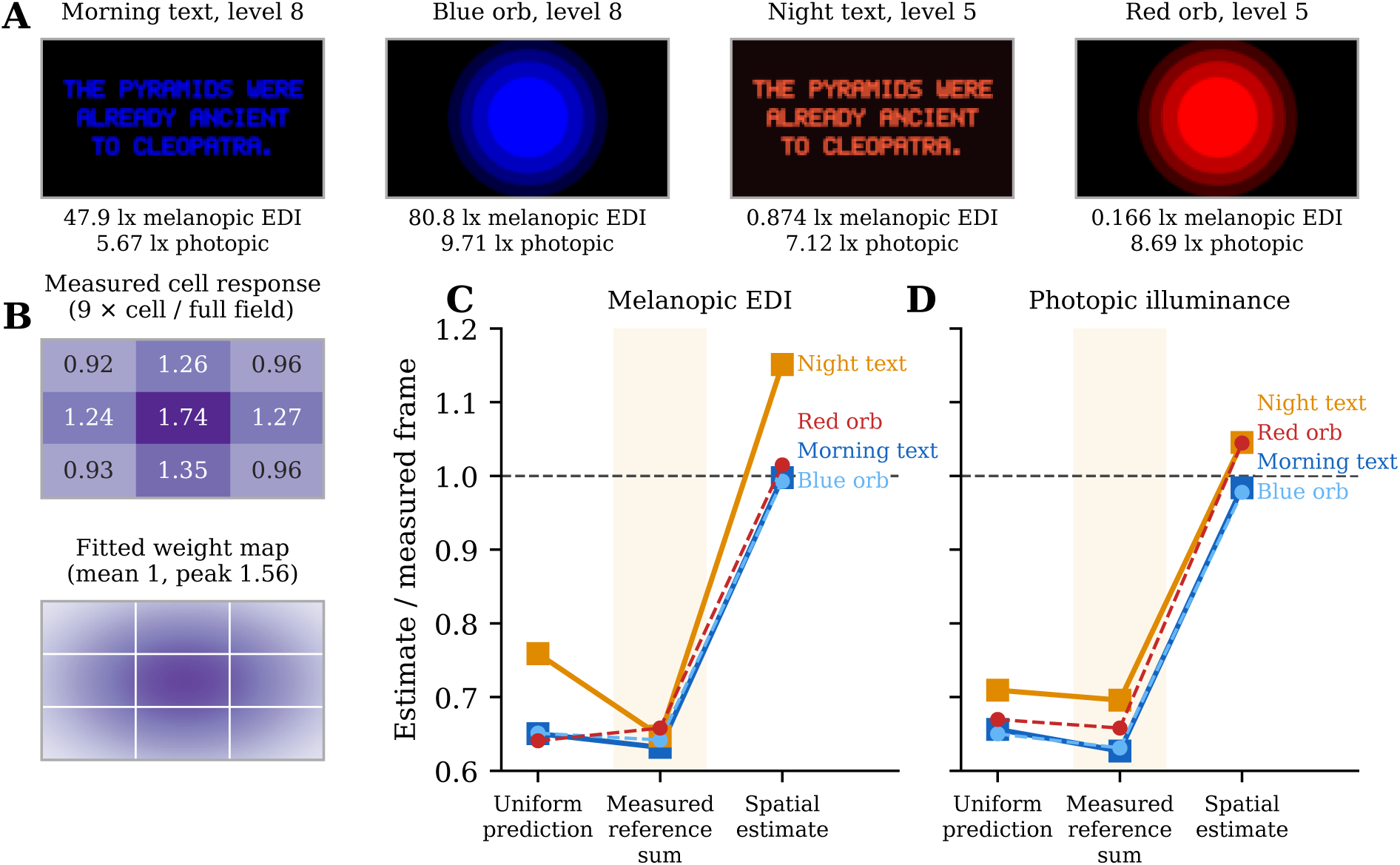
Limits of uniform-field calibration for rendered application content. (**A**) The four example frames (morning text slide, blue orb, night text slide in the mockup application’s colors, and red orb), as shown on the glasses (downsampled), with the measured melanopic EDI and photopic illuminance of each frame (mean of 3 sessions) below it. (**B**) Spatial grid. Top: photopic illuminance of each of nine equal cells lit alone at full code, times nine, over that of the full field (mean of red, green, blue, and white and of 3 sessions at level 7); 1 everywhere would be equal weighting, and the values also include the lit-area gain. Bottom: the fitted spatial weight map, normalized to mean 1 over the frame, with the grid cells outlined. (**C**, **D**) Transfer of the uniform-field calibration to rendered frames, for (**C**) melanopic EDI and (**D**) photopic illuminance, one line per frame (text slides solid with squares, orbs dashed with circles). Ratios are each estimate divided by the measured frame exposure (mean of 3 sessions; 1 is exact agreement, dashed line): the uniform prediction from the locked model with equal spatial weighting; the pixel-weighted sum of the same-session measured uniform references (shaded), which involves no model; and the spatial correction model derived independently from the 3 *×* 3 grid. Uniform predictions and measured uniform-reference sums are similarly low, showing that the discrepancy arises from the rendered frame’s spatial behavior rather than primarily from model error; the spatial correction largely removes it. Absolute values are in Table 13.

The spatial grid accounts for this excess with two effects (Fig. 5B). First, the measurement geometry is more sensitive to the center of the displayed field: the cell response, nine times a single cell over the full field, which also includes the lit-area gain, was 1.65–1.87 for the center cell, 1.19–1.46 for the side cells, and 0.87–1.04 for the corners (photopic, mean of the 3 sessions and four colors), and the fitted weight map peaks at 1.56 times its mean. All four frames center their bright content. Second, a small lit area gives more light per pixel than a full field: the nine cells summed to 1.14–1.15 times the full field for the primaries and 1.28 for white, rather than 1, consistent with display-level power limiting, in which per-pixel output decreases as a larger fraction of the display is illuminated (fitted lit-area exponents in Appendix A.1). With both terms, a secondary spatial correction model, derived from this independent grid, accounted for position-dependent weighting and lit-area gain. Applied to the rendered frames without fitting to those frame measurements, it reduced the discrepancy between predicted and measured exposure substantially (Fig. 5C,D): the corrected estimates differed from the measured frames by 0%, -1%, +15%, and +1% for melanopic EDI (photopic illuminance in Table 13). Because the four frames covered only a narrow range of illuminated area and intensity and were displayed at brightness levels different from the spatialgrid measurement, this agreement was evaluated as a consistency test rather than an independently validated prediction model.

## 4 Discussion

### 4.1 From consumer XR display to calibrated photostimulation platform

This study shows that a consumer see-through XR display can be converted into a quantitatively controlled ocular-light platform, but only by measuring the relationship between requested display settings and light delivered at the eye. The central contribution is therefore not a characterization of one pair of glasses, but a workflow that links control, measurement, calibration, held-out validation, and explicitly defined conditions under which the calibration applies. The requested code and brightness level do not specify the light at the eye: important display behaviors that set the delivered exposure appear in no manufacturer specification (Section 4.2), and rendered frames depart further still from their uniform parts. With purpose-built control software, measurement at the corneal plane, and a model whose error is quantified on held-out settings (Section 3.4), the same device spans light-poor to daytime-level melanopic stimulation along three independently controlled and logged intensity variables (RGB code, hardware brightness, and duty cycle), with a spectrally stable blue primary, reseating repeatability of a few percent, and blue-light-hazard exposure that remained well below the conservative screening limit [26, 27]. Together, these steps provide a workflow that can be adapted to characterize other head-worn displays.

Prior head-mounted work has already calibrated an immersive VR headset in detail and shown that its light suppresses melatonin [17]. The distinction here is see-through operation, together with independently controlled and logged hardware states and a calibration workflow that can be reapplied to other head-worn displays [22]. Full occlusion removes stable real-world visual references and is associated with cybersickness and related adverse effects [18]; see-through glasses may preserve those references while keeping the source fixed relative to the head. Whether this confers a comfort or usability advantage was not tested. In supervised settings such as the intensive care unit, see-through operation could facilitate monitored administration of a logged stimulus, but wearer comfort, the visibility of the patient’s eyes to caregivers, and clinical usability were not evaluated.

The trade-off is ambient light. An unmodified see-through form factor does not exclude it as completely as an occlusive headset: the electrochromic film reduces but does not block peripheral light, so ambient exposure adds to the display stimulus and remains a covariate. Protocols that need near-exclusive control of retinal input can add light-isolation accessories.

### 4.2 What the device characterization means for experimental control

Several of the measured properties change how an experiment on this device should be designed. Green is not a low-melanopic control: at the same level it delivers as much melanopic EDI as blue, although its melanopic DER is much lower, so the two differ mainly in photopic illuminance and cone-mediated visual stimulation. White is less than the sum of its primaries, with the shortfall growing with output, so mixed stimuli must be measured, or predicted by a model calibrated on measured mixtures (Section 3.4), and white cannot be run at the top brightness level at all. The brightness steps are uneven and the code curve departs reproducibly from a power law, so both are best used as measured lookup tables; a protocol that assumed additive mixtures, even brightness steps, or an exact power law in code would mis-specify the stimulus, in some cases by more than a factor of two. Logging the controllable hardware states relevant to intensity with each capture allows a session record to be checked against a measured calibration, and the between-session spread of a few percent sets how closely that calibration predicts a new fitting of the glasses.

The full-color panel also allows the spectrum to be scheduled. The blue and red primaries differ 467-fold in melanopic DER, so the same worn device can deliver blue-weighted daytime stimulation and red-shifted evening illumination, where self-luminous displays used in the evening otherwise delay circadian phase and suppress melatonin [28]. At every brightness level the red channel stays well below the 10 lx pre-sleep maximum for healthy adults [2], although use during a dark sleep period would require a low level, a lower code, or a reduced duty cycle. A blue-versus-red comparison produces a large melanopic contrast but also changes cone stimulation, and matched visual salience was not tested. Because code, hardware brightness, and duty cycle are set and logged independently, spectral composition can be manipulated independently of overall light intensity within a subject rather than across lighting rigs. The red primary’s low melanopic output could also carry visual instructions or other interactive content when melanopic exposure should be minimized, subject to calibration of the rendered content (Section 4.3) and to ambient exposure.

### 4.3 From calibrated settings to application design

Within its calibrated domain, uniform full-field stimuli, the settings-to-exposure model (Sections 2.5 and 3.4) turns the calibration into a design tool. Software can choose a uniform color, brightness level, and duty cycle and obtain the predicted corneal melanopic EDI, photopic illuminance, and melanopic DER from the full-precision calibration package without a new measurement, for example to hold melanopic EDI constant while changing color, to step it on a schedule, or to keep a uniform evening stimulus below a specified maximum melanopic EDI. Each prediction carries a status (valid, low signal, unsupported, or configuration mismatch) and states whether the setting was measured, interpolated, or extrapolated, so software can reject unsupported settings and identify predictions that require low-code extrapolation (Appendix A).

Rendered content lies outside this validated domain (Sections 2.6 and 3.5), and uniform prediction fails for structured frames: nonlinear pixel values make mean RGB inadequate, and even the pixel-weighted sum of uniform predictions was low by about a third in melanopic EDI. Independent spatial measurements then showed why, because exposure depends on where and how much of the display is illuminated: the measurement geometry weights the center of the field, and a small lit area gives more light per pixel than a full field. A secondary spatial correction model, derived from an independent 3 *×* 3 display grid and applied to the rendered frames without fitting to those frame measurements, accounted for both effects and substantially restored agreement, coming within a few percent of the measured frames except for the night slide’s melanopic EDI (+15%, under 1 lx). Together, these results show that rendered-frame exposure can be estimated more accurately by accounting for both spatial weighting and illuminated-area gain, providing a practical extension of the uniform-field calibration to application-level content. The repository’s developer usage guide documents the corresponding composition rules (Data availability).

### 4.4 Biological and clinical research applications

At its top usable settings the device reaches daytime-level stimulation, and its range down the brightness and code axes spans the levels at which melatonin suppression is graded and the levels measured in hospital care units (Fig. 6) [11, 15, 16]. In a unit whose daytime melanopic EDI averages 55 lx [15], a worn source adding up to 269 lx of logged, spectrally fixed melanopic EDI would be a substantial and controllable increment that does not depend on window orientation or bed position. The fine control at the low end also suits evening and exposure–response protocols, in which the stimulus must be set below or near suppression thresholds. These are research applications; no physiological or clinical effect of this device was tested.

**Figure 6:**
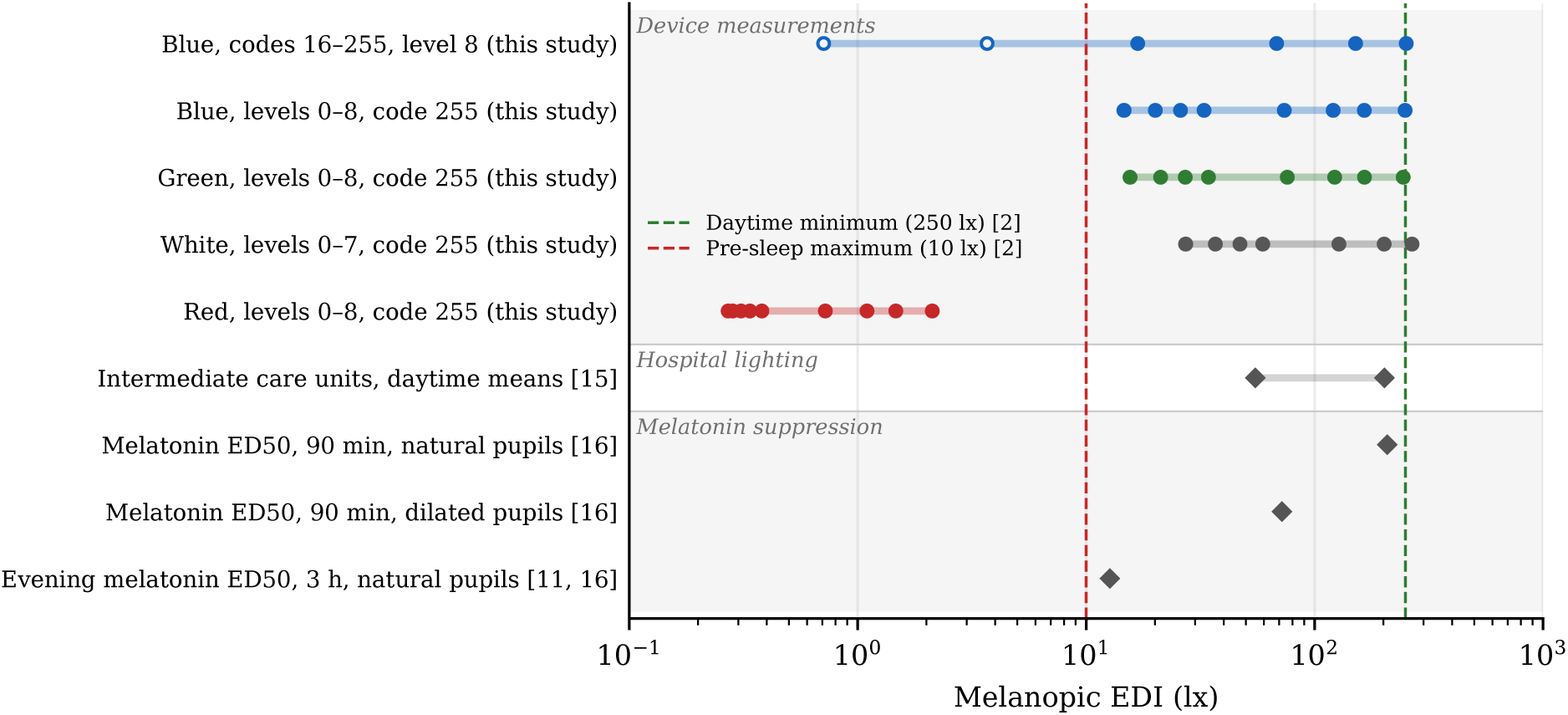
Measured corneal melanopic EDI against published reference levels. Rows are grouped into three blocks: device measurements (circles, means of three sessions; what the glasses deliver: the blue code series and the brightness series of each channel, with their range; open circles are values whose photopic illuminance is below the meter’s specified 1 lx minimum and are approximate; that minimum is photopic and does not correspond to 1 lx on this melanopic axis, so no line is drawn for it), hospital lighting (the environments the glasses could supplement), and melatonin-suppression benchmarks (evidence that responses occur within this general exposure range). Diamonds and dashed lines: published values, with reference numbers on the axis [2, 11, 15, 16]. The hospital row spans the daytime (06:00–17:00) mean melanopic EDI of two intermediate care units, 55 and 202 lx [15], recorded by a detector behind the bed rather than at the eye. The evening melatonin benchmark [11] refers to a three-hour assessment window, whereas the pooled estimates [16] refer to 90-min exposures. The published values were obtained under different exposure conditions (room-level sensors, full-field or room-like exposures, healthy adults) and are contextual estimates, not predicted responses to this display.

**Figure 7:**
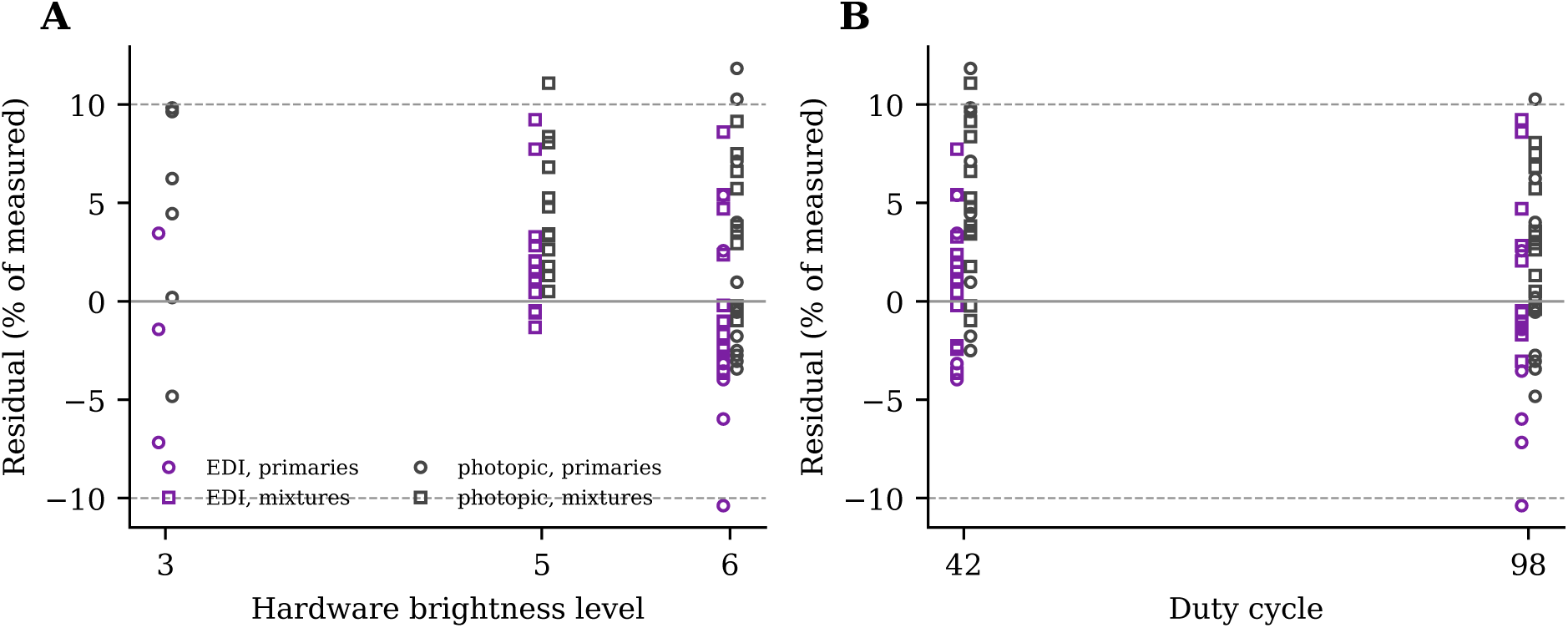
Validation residuals of the settings-to-exposure model. Percentage residuals (predicted minus measured) of melanopic EDI (purple) and photopic illuminance (gray) for single-primary (circles) and mixed (squares) stimuli eligible for that output, against (**A**) hardware brightness level and (**B**) duty cycle, offset slightly for visibility; dashed lines at *±*10%.

Two cautions apply to the comparison. The display lights a central portion of the visual field, and equal corneal melanopic EDI from a central portion may not act like a full-field room exposure, from which the exposure–response estimates largely come [16]. And the hospital values were recorded with a spectral detector behind the bed at 1.15 m, angled to approximate the light reaching the patient’s eyes but not at eye level [15].

Controlled ocular exposure is also relevant beyond circadian timing. Preclinical work links short-wavelength ocular exposure to autonomic and immune outcomes in models of critical illness, through a pathway that requires an intact optic nerve [19–21], and light has begun to be tested in perioperative and intensive-care settings [20, 29], where the light at the eye was not controlled. A worn source with a logged spectrum and intensity would let mechanistic studies vary the ocular stimulus deliberately and hold it constant across patient positions.

### 4.5 Scope and limitations

#### Device and generalizability

The results come from one unit, measured in three sessions within 22 min on one day at the right eye position with both displays lit, on a rigid phantom. Repeatability was assessed over successive fittings of the glasses, not over days, units, or wearers, and the between-session spread of a few percent describes reseating on the phantom rather than fit on a human head; absolute values describe this geometry, and output stability over longer exposures was not measured. The meter’s specifications bound the absolute values: illuminance accuracy is specified for illuminant A, not for narrowband sources; its spectral bandwidth broadens the measured spectra; and the dimmest blue codes fall below its specified 1 lx photopic minimum (Section 3.3) [25]. Full white could not be run at the top brightness level on either host platform tested, and the cause within the device was not investigated.

#### Exposure interpretation

All quantities are corneal exposure intensities: pupil size, ocular media, ambient light, and exposure duration, which set the retinal stimulus and dose, were not modeled. If the specified 52^◦^ field of view is diagonal, the display lights a central field of about 45^◦^*×*29^◦^.

#### Model domain

The settings-to-exposure model was calibrated and validated only on uniform full-frame stimuli, on this unit, in this geometry, with the film off and the display mode used here; a prediction for a different configuration is flagged rather than corrected, and it does not extrapolate beyond the calibrated levels and duty cycles. Its remaining caveats are given in Appendix A.1. The spatial correction was evaluated as a consistency test rather than an independently validated prediction model: its weight map describes the phantom and probe, not a wearer’s eye, and it was formulated after the frames were measured and has not been tested on a new set of rendered frames. Time-varying content was not tested.

These limitations define the scope of the present calibration rather than the performance of the device outside the tested conditions.

## 5 Conclusion

Consumer see-through XR displays can be converted from entertainment devices into quantitatively controlled ocular-light platforms by linking software state to direct corneal-plane measurement. In the tested device, this workflow provided spectrally stable, independently controlled light spanning light-poor to daytime-relevant melanopic exposures, while a calibration model predicted held-out uniform stimuli with mean absolute percentage errors of 3.4% for melanopic EDI and 4.5% for photopic illuminance. Rendered application frames revealed position- and illuminated-area-dependent exposure, and a spatial correction derived from an independent display grid substantially improved agreement with measured frame exposure. Together, these results provide a reproducible framework for developing and evaluating head-worn photostimulation tools for circadian, neurobiological, and clinical research.

## Data Availability

All data produced in the present study are available upon reasonable request to the authors.

https://github.com/mattgaidica/XR-Glasses-Characterization

## Acknowledgments

We thank the Washington University Neurotech Hub for fabrication expertise and resources.

## Author contributions

M.G. designed and built the stimulus host application, the phantom-head measurement fixture, and the acquisition and analysis software, performed the measurements, and drafted the manuscript. M.R. contributed to study framing and clinical interpretation. Both authors reviewed and approved the final manuscript.

## Data availability

The characterization software (stimulus host application and acquisition and analysis package), the 3D model of the spectroradiometer used to build the phantom fixture, the reference estimator, the calibration package of the settings-to-exposure model (luma_ultra_v1; parameters in Table 10) with its validation cases, a developer usage guide, and the data underlying the results presented in this paper (sessions 20261004_114931, 20261004_115834, and 20261004_120607: raw spectra, capture metadata, per-capture analysis outputs, and the host event logs) are available under the MIT License at https://github.com/mattgaidica/XR-Glasses-Characterization. The scripts that generate every figure, table, and numeric value in this manuscript are kept with the manuscript source and are not part of that distribution. The version described here is commit 8763fdd7b22d5bc8dde409896d9f9c3f8f4dea93 of that repository. The phantom head was built from third-party models that cannot be redistributed; the repository identifies them and gives the geometry needed to rebuild the fixture.

## A Implementing the settings-to-exposure model

This appendix specifies the algorithm of the settings-to-exposure model of Section 2.5 completely enough to implement it in any language. Reproducing the predictions of Section 3.4 exactly requires the full-precision parameters in the JSON calibration package (calibration luma_ultra_v1; check-sum below) and, for a crossed model, its grid-node data; Table 10 gives the parameters rounded for reading. The reference implementation is the estimator in the analysis package. The inputs are a uniform full-frame code (*k_R_, k_G_, k_B_*) with integer values 0–255, a hardware brightness level *L ∈ {*0*, . . . ,* 8*}*, and a duty cycle *D* in percent. Melanopic EDI and photopic illuminance are computed by the same steps, each with its own parameters; both are display-attributable corneal-plane values in lux, and neither includes ambient light.

### Step 0: configuration

The package records the device, firmware, product identifier, film state, and display mode it was calibrated in. If the caller supplies the current configuration and any of these differs, the status is configuration mismatch and no value is returned. The package covers uniform full-frame stimuli only.

### Step 1: shared input restrictions

These apply to the request as a whole: if any fails, the status is unsupported and neither output is returned. The stimulus must not have more than one nonzero channel at *L >* 7 (this includes white) or at *L* below the lowest level at which the mixtures were calibrated (4; recorded in the package), and Steps 2–4 must give a photopic value, which requires *L* to be a calibrated level of every nonzero channel, *D* to lie within the calibrated duty range (30–98), and, for a code below 255, *L* to lie within the channel’s code-series levels *L*_1_ *. . . L_n_*, the levels at which its code nodes were fitted (code 0, the channel off, is defined at every level). The photopic parameters define this shared domain. Missing support in the melanopic parameters alone does not reject the request; it is handled per output in Step 6. The only extrapolation is in code, below a level’s lowest node (Step 2).

### Step 2: code response

At code-series level *L_j_*a channel has code nodes *k*_1_ *< · · · < k_m_* below 255 with values *a*_1_*, . . . , a_m_*; set *k_m_*_+1_ = 255 and *a_m_*_+1_ = 1. For *k_i_ ≤ k ≤ k_i_*_+1_,

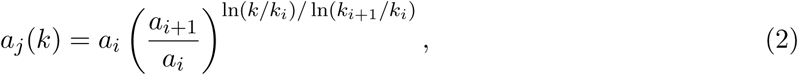

and for 0 *< k < k*_1_ the same expression with *i* = 1 (the lowest segment extended toward code 0; the result is reported as extrapolated). For *L_j_ < L < L_j_*_+1_, with *t* = (*L − L_j_*)*/*(*L_j_*_+1_ *− L_j_*),

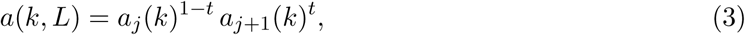

so that *a*(255*, L*) = 1 at every level and *a*(0*, L*) = 0.

### Step 3: duty response

With calibrated duty cycles *D*_1_ *< · · · < D_m_* and values *q*_1_*, . . . , q_m_* (*q_m_* = 1), for *D_j_ ≤ D ≤ D_j_*_+1_,

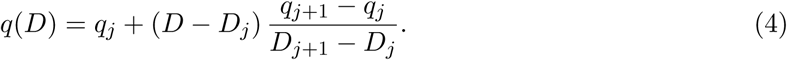

### Step 4: primaries

Each nonzero channel contributes *y_c_* = *a_c_*(*k_c_, L*) *b_c_*(*L*) *q_c_*(*D*), where *b_c_*(*L*) is the channel’s full-code output at level *L* and the highest calibrated duty cycle.

### Step 5: mixture

With *g* = *β*_0_ + *β_L_L/*8 + *β_D_D/*100, the prediction is, for the total form of the interaction term,

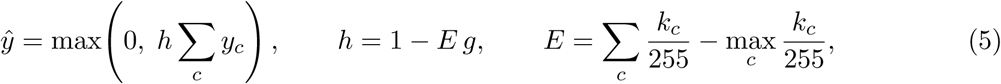

and for the per-channel form

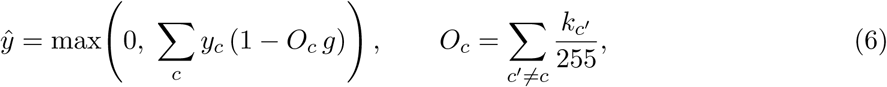

so that a single channel is unchanged in both forms; *g* = 0 throughout if the selected model has no interaction term. The package records which form the model uses.

### Step 6: DER and status, per output

Each output’s parameters cover only settings whose calibration measurements passed that output’s signal test (Section 2.5), so the melanopic domain can be smaller than the photopic one. If Steps 1–5 give a photopic value but the melanopic parameters do not cover the setting (its code, level, or duty lies outside them), the status is low signal: photopic illuminance is returned, and melanopic EDI and the DER are not. If the predicted photopic illuminance is below 1 lx, the meter’s specified photopic minimum, both values are returned with a low-signal status and no DER, because the setting lies below the photopic range in which the model was evaluated; a melanopic EDI below 1 lx does not by itself trigger this status. Otherwise the melanopic DER is *y*^_EDI_*/y*^_lux_ and the status is valid. A setting is reported as measured if it is a calibration setting, as extrapolated if any code lies below its channel’s lowest node at that level (Step 2), and as interpolated otherwise. The reference software also attaches a warning when the package’s model is outside the fitting tool’s default development thresholds (MAPE *≤* 5% and P95 *≤* 10%); these are engineering defaults of the software, not criteria of this evaluation.

### Crossed models

If the selected model is crossed, Step 4 uses the measured grid instead of *a_c_* and *q_c_*: at a grid level, the measured outputs *v_i_* at codes *k_i_ ≤ k ≤ k_i_*_+1_ are interpolated in code by the power-law segment *v*(*k*) = *v_i_* (*v_i_*_+1_*/v_i_*)^ln(*k/k*^*^i^*^)*/* ln(*k*^*^i^*^+1^*^/ki^*^)^ (linear if either value is not positive) at the two calibrated duty cycles bracketing *D* (or at *D* itself if it is calibrated), and the results are interpolated in duty as in Step 3. Only these bracketing nodes need contain the code; other duty nodes at that level may lack it (their settings were not eligible) without making the setting unsupported. At other levels, the value at the nearest grid level *L*^∗^ (the higher one if two are equally near) is multiplied by *b_c_*(*L*)*/b_c_*(*L*^∗^). The grid nodes are in the calibration package.

### Rendered content

The estimator above covers uniform full-frame stimuli only. For rendered frames the analysis package also reports two quantities outside that validated estimator: the pixel-weighted sum of uniform estimates (equal spatial weighting), and a secondary spatial correction (Section 2.6), which multiplies each color’s uniform estimate by the mean of the grid’s weight map over its pixels and by the lit-area gain ratio (load_frame_*/*load_color_)^−^*^α^* (Section 3.5). The correction was derived from the independent grid and applied without fitting to the frame measurements; it was evaluated as a consistency test rather than an independently validated prediction model (Section 4.5).

### Worked example

For (192, 96, 0) at level 6 and duty 42, level 6 lies between the code-series levels 5 and 7, so each *a_c_*(*k_c_,* 6) is interpolated log-linearly between them (Step 2), and duty 42 is a calibrated duty cycle (Step 3). For melanopic EDI, *y_R_* = 0.4258 *×* 1.019 *×* 0.4548 = 0.1974; *y_G_* = 0.07520*×*122.3*×*0.4494 = 4.135; their sum is 4.332 lx; *g* = 0.07211, *h_R_* = 0.9729, *h_G_* = 0.9457; so the prediction is Σ*_c_ h_c_y_c_* = 4.102 lx. For photopic illuminance, *y_R_* = 0.4258 *×* 55.73 *×* 0.4548 = 10.79; *y_G_* = 0.07823 *×* 198.4 *×* 0.4493 = 6.973; their sum is 17.76 lx; *g* = 0.06377, *h_R_* = 0.9760, *h_G_*= 0.9520; so the prediction is Σ*_c_ h_c_y_c_* = 17.17 lx. The melanopic DER is 0.239.

### Reproducibility

The calibration and validation grids (grid version v3 and v3-topup) are listed in Table 5, the cross-validation comparison of the seven candidates, with and without anchor normalization, in Table 6, and the validation error by composition class, which tests the interaction term across channel pairs, in Table 8. Table 10 is rounded for reading and is not sufficient on its own to reproduce the predictions; an implementation should load the full-precision calibration package (data/uprtek/model/calibration.json, SHA-256 a592af30d04ed83e7b9e6996d1d6d6ee eebf2f7304fc2569a7435a06ba6f422a), including its grid nodes for a crossed model, and should reproduce the reference cases of Table 9, which were computed by the reference estimator from that package.

**Table 5:** Calibration and validation grids. Each setting is a uniform full-frame code at every listed level and duty cycle. *n*: settings per session.

| Stage | Stimuli (R, G, B) | Levels | Duty cycles | $n$ |
| --- | --- | --- | --- | --- |
| Primaries (calibration) | R, G, and B alone at codes 64 and 192 | 8 | 98 | 6 |
|  | R and G alone at codes 128 and 255 | 4 and 8 | 30, 42, and 98 | 24 |
|  | B alone at codes 128 and 255 | 8 | 30, 42, and 98 | 6 |
|  | B alone at code 255 | 4 | 30, 42, and 98 | 3 |
| Ladders (calibration) | R, G, and B alone at codes 64, 128, and 192 | 2, 4, 5, and 7 | 98 | 36 |
| Mixtures (calibration) | (64, 64, 64), (128, 128, 128), (192, 192, 192), (255, 255, 255), (128, 128, 0), (128, 0, 128), (0, 128, 128), (255, 255, 0), (255, 0, 255), (0, 255, 255), (255, 64, 0), (64, 255, 0), (0, 255, 64), (0, 64, 255), (255, 0, 64), (64, 0, 255), (255, 128, 64), (64, 255, 128), (128, 64, 255) | 4 and 7 | 30 and 98 | 76 |
| Supplemental mixtures (calibration) | (255, 64, 0), (64, 255, 0), (0, 255, 64), (0, 64, 255), (255, 0, 64), (64, 0, 255), (64, 64, 64), (128, 128, 128) | 6 | 98 | 8 |
| Validation primaries | R, G, and B alone at codes 96 and 160 | 3 and 6 | 42 and 98 | 24 |
| Validation mixtures | (96, 96, 96), (160, 160, 160), (192, 96, 0), (0, 192, 96), (96, 0, 192), (192, 96, 80) | 5 and 6 | 42 and 98 | 24 |

**Table 6:** Model candidates compared by leave-one-session-out cross-validation on the calibration sessions. The candidate with the fewest failed predictions and then the lowest worst-case MAPE is selected (the fitting software first prefers the simplest candidate within its default development thresholds, MAPE *≤* 5% and P95 *≤* 10% for both outputs with no failures; no candidate was within them). +H: total interaction term; +Hc: per-channel interaction term. Normalized: MAPE of the same cross-validation with every calibration session scaled by its blue anchor, reported for comparison and not used for selection.

| Candidate | EDI (%) |  | Photopic (%) |  | Failed | Normalized (%) |  |  |
| --- | --- | --- | --- | --- | --- | --- | --- | --- |
|  | MAPE | P95 | MAPE | P95 |  | EDI | Phot. |  |
| factorized | 7.2 | 26.8 | 6.1 | 18.2 | 33 | 6.7 | 5.4 |  |
| factorized+H | 5.9 | 21.7 | 5.0 | 14.6 | 33 | 5.2 | 4.0 |  |
| factorized+Hc | 5.5 | 17.2 | 4.9 | 14.0 | 33 | 5.0 | 3.9 |  |
| factorized+Hc+Rder | 6.1 | 20.0 | 4.9 | 14.0 | 0 | 5.5 | 3.9 | selected |
| crossed | 4.8 | 15.7 | 5.1 | 16.3 | 203 | 4.2 | 4.5 |  |
| crossed+H | 3.3 | 10.8 | 3.8 | 12.4 | 203 | 2.4 | 2.8 |  |
| crossed+Hc | 3.4 | 10.8 | 3.8 | 12.4 | 203 | 2.4 | 2.8 |  |

**Table 7:** Prediction error of the selected model by group (single primaries and mixtures), on the independent validation session and in cross-validation; columns as in Table 4.

| Output | Stimuli | Independent validation |  |  |  |  | Cross-validation |
| --- | --- | --- | --- | --- | --- | --- | --- |
| | | $n$ | MAPE (%) | P95 (%) | Bias (%) | MAE (lx) | MAPE (%) |
| Melanopic EDI | All | 34 | 3.4 | 8.8 | 0.3 | 0.39 | 6.1 |
|  | Primaries | 10 | 4.7 | 8.9 | -2.4 | 0.35 | 2.9 |
|  | Mixtures | 24 | 2.9 | 8.5 | 1.4 | 0.43 | 8.0 |
| Photopic | All | 41 | 4.5 | 10.3 | 3.5 | 0.38 | 4.9 |
|  | Primaries | 17 | 4.9 | 10.6 | 2.7 | 0.22 | 4.4 |
|  | Mixtures | 24 | 4.3 | 9.0 | 4.1 | 0.54 | 5.4 |
| Melanopic DER | All | 34 | 3.5 | 8.6 | -2.4 | — | 4.5 |
|  | Primaries | 10 | 2.2 | 5.3 | -2.2 | — | 1.6 |
|  | Mixtures | 24 | 4.0 | 8.6 | -2.5 | — | 6.2 |

**Table 8:** Validation error of the selected model by composition class (settings eligible for each output).

| Composition | Melanopic EDI |  |  | Photopic |  |  |
| --- | --- | --- | --- | --- | --- | --- |
| | $n$ | MAPE (%) | Bias (%) | $n$ | MAPE (%) | Bias (%) |
| red alone | 0 | — | — | 6 | 6.6 | 6.4 |
| green alone | 7 | 3.8 | -0.6 | 8 | 4.4 | 1.9 |
| blue alone | 3 | 6.8 | -6.8 | 3 | 2.8 | -2.8 |
| gray ($R = G = B$) | 8 | 1.9 | -0.0 | 8 | 5.1 | 5.0 |
| red + green | 4 | 7.7 | 7.7 | 4 | 4.0 | 4.0 |
| red + blue | 4 | 2.0 | -1.0 | 4 | 6.9 | 6.9 |
| green + blue | 4 | 1.1 | -0.9 | 4 | 0.9 | 0.2 |
| three channels, unequal | 4 | 2.6 | 2.6 | 4 | 3.5 | 3.5 |

**Table 9:** Reference input/output cases computed by the reference estimator from the calibration package (luma_ultra_v1). Duty cycles are taken from the package’s calibrated duty cycles (the highest, the middle one for the calibration setting, the lowest for the dim primary, and the midpoint of the two highest for interpolated cases), so the cases remain measured, interpolated, or in domain under either duty protocol. Values are rounded to four significant figures; – marks a value that is not returned.

| Case | $(R, G, B), L, D$ | EDI (lx) | Photopic (lx) | DER | Status | Support |
| --- | --- | --- | --- | --- | --- | --- |
| Interpolated primary | (0, 0, 160), 6, 42 | 15.45 | 1.883 | 8.207 | ok | interpolated |
| Mixture (example) | (192, 96, 0), 6, 42 | 4.102 | 17.17 | 0.2389 | ok | interpolated |
| Calibration setting | (0, 0, 128), 8, 42 | 30.44 | 3.679 | 8.273 | ok | measured |
| Dim primary | (64, 0, 0), 2, 30 | 0.001647 | 0.09005 | – | low signal | extrapolated |
| Code below 255 under $L_1$ | (128, 0, 0), 1, 98 | – | – | – | unsupported | unsupported |
| Code below $k_1$ | (0, 0, 32), 6, 42 | 0.2323 | 0.03786 | – | low signal | extrapolated |
| White at level 8 | (255, 255, 255), 8, 98 | – | – | – | unsupported | unsupported |
| Firmware differs | (0, 0, 255), 8, 98 | – | – | – | configuration mismatch | unsupported |

**Table 10:**
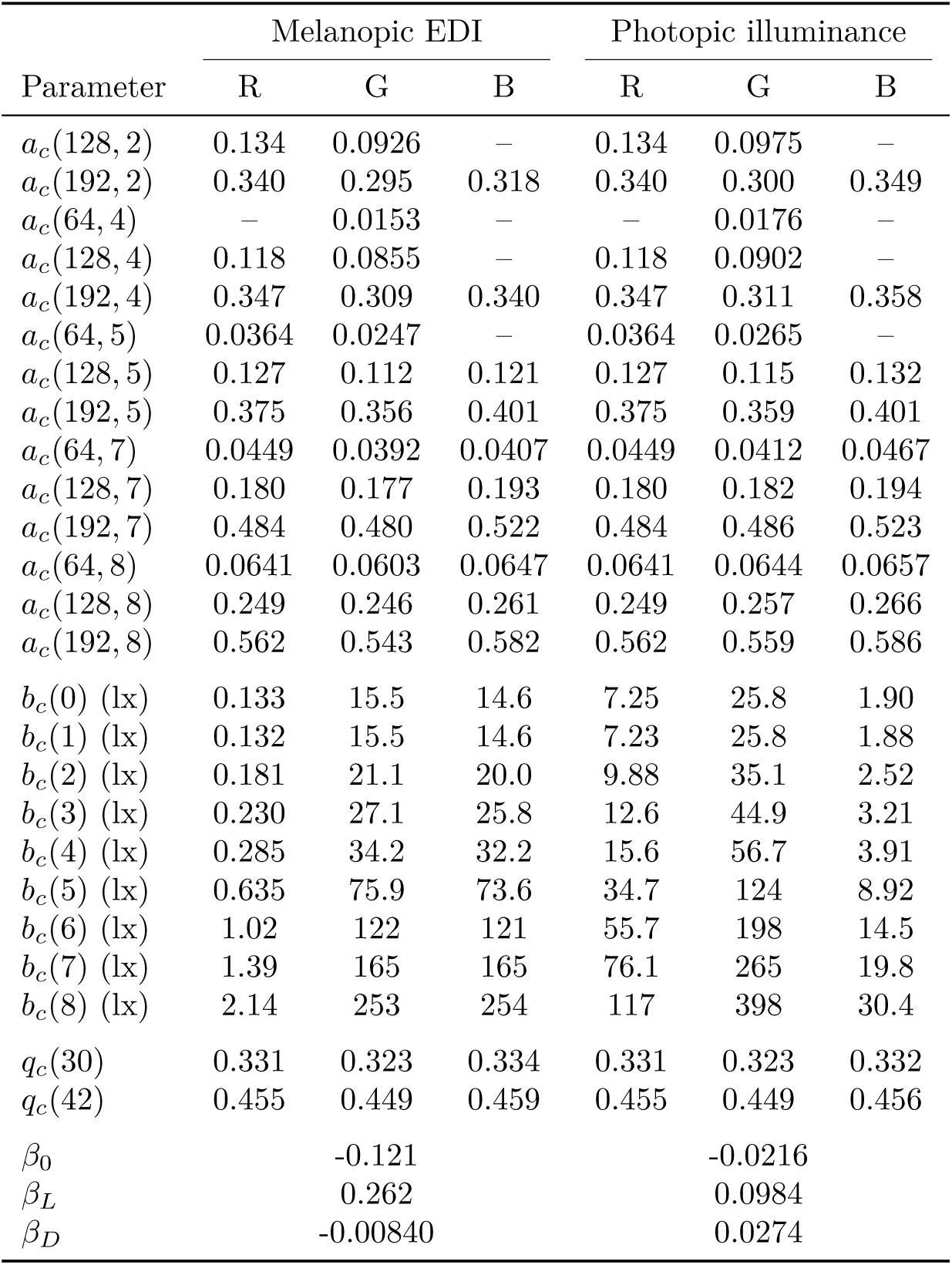
Parameters of the settings-to-exposure model (calibration luma_ultra_v1), rounded for reading; the full-precision values are in the calibration package (SHA-256 a592af30d04ed83e7b9e6996d1d6d6ee eebf2f7304fc2569a7435a06ba6f422a). *a_c_*(*k, L*): code node, the output at code *k* relative to code 255 at code-series level *L* (Step 2); *b_c_*(*L*): full-code output at level *L* and the highest calibrated duty cycle (98); *q_c_*(*D*): duty response relative to that duty cycle; *β*: mixture interaction coefficients (Step 5). By definition *q_c_*(98) = 1; dashes mark parameters that the selected model does not use.

| Parameter | Melanopic EDI |  |  | Photopic illuminance |  |  |
| --- | --- | --- | --- | --- | --- | --- |
|  | R | G | B | R | G | B |
| $a_c(128, 2)$ | 0.134 | 0.0926 | – | 0.134 | 0.0975 | – |
| $a_c(192, 2)$ | 0.340 | 0.295 | 0.318 | 0.340 | 0.300 | 0.349 |
| $a_c(64, 4)$ | – | 0.0153 | – | – | 0.0176 | – |
| $a_c(128, 4)$ | 0.118 | 0.0855 | – | 0.118 | 0.0902 | – |
| $a_c(192, 4)$ | 0.347 | 0.309 | 0.340 | 0.347 | 0.311 | 0.358 |
| $a_c(64, 5)$ | 0.0364 | 0.0247 | – | 0.0364 | 0.0265 | – |
| $a_c(128, 5)$ | 0.127 | 0.112 | 0.121 | 0.127 | 0.115 | 0.132 |
| $a_c(192, 5)$ | 0.375 | 0.356 | 0.401 | 0.375 | 0.359 | 0.401 |
| $a_c(64, 7)$ | 0.0449 | 0.0392 | 0.0407 | 0.0449 | 0.0412 | 0.0467 |
| $a_c(128, 7)$ | 0.180 | 0.177 | 0.193 | 0.180 | 0.182 | 0.194 |
| $a_c(192, 7)$ | 0.484 | 0.480 | 0.522 | 0.484 | 0.486 | 0.523 |
| $a_c(64, 8)$ | 0.0641 | 0.0603 | 0.0647 | 0.0641 | 0.0644 | 0.0657 |
| $a_c(128, 8)$ | 0.249 | 0.246 | 0.261 | 0.249 | 0.257 | 0.266 |
| $a_c(192, 8)$ | 0.562 | 0.543 | 0.582 | 0.562 | 0.559 | 0.586 |
| $b_c(0)$ (lx) | 0.133 | 15.5 | 14.6 | 7.25 | 25.8 | 1.90 |
| $b_c(1)$ (lx) | 0.132 | 15.5 | 14.6 | 7.23 | 25.8 | 1.88 |
| $b_c(2)$ (lx) | 0.181 | 21.1 | 20.0 | 9.88 | 35.1 | 2.52 |
| $b_c(3)$ (lx) | 0.230 | 27.1 | 25.8 | 12.6 | 44.9 | 3.21 |
| $b_c(4)$ (lx) | 0.285 | 34.2 | 32.2 | 15.6 | 56.7 | 3.91 |
| $b_c(5)$ (lx) | 0.635 | 75.9 | 73.6 | 34.7 | 124 | 8.92 |
| $b_c(6)$ (lx) | 1.02 | 122 | 121 | 55.7 | 198 | 14.5 |
| $b_c(7)$ (lx) | 1.39 | 165 | 165 | 76.1 | 265 | 19.8 |
| $b_c(8)$ (lx) | 2.14 | 253 | 254 | 117 | 398 | 30.4 |
| $q_c(30)$ | 0.331 | 0.323 | 0.334 | 0.331 | 0.323 | 0.332 |
| $q_c(42)$ | 0.455 | 0.449 | 0.459 | 0.455 | 0.449 | 0.456 |
| $\beta_0$ | | -0.121 | | | -0.0216 | |
| $\beta_L$ | | 0.262 | | | 0.0984 | |
| $\beta_D$ | | -0.00840 | | | 0.0274 | |

**Table 11:**
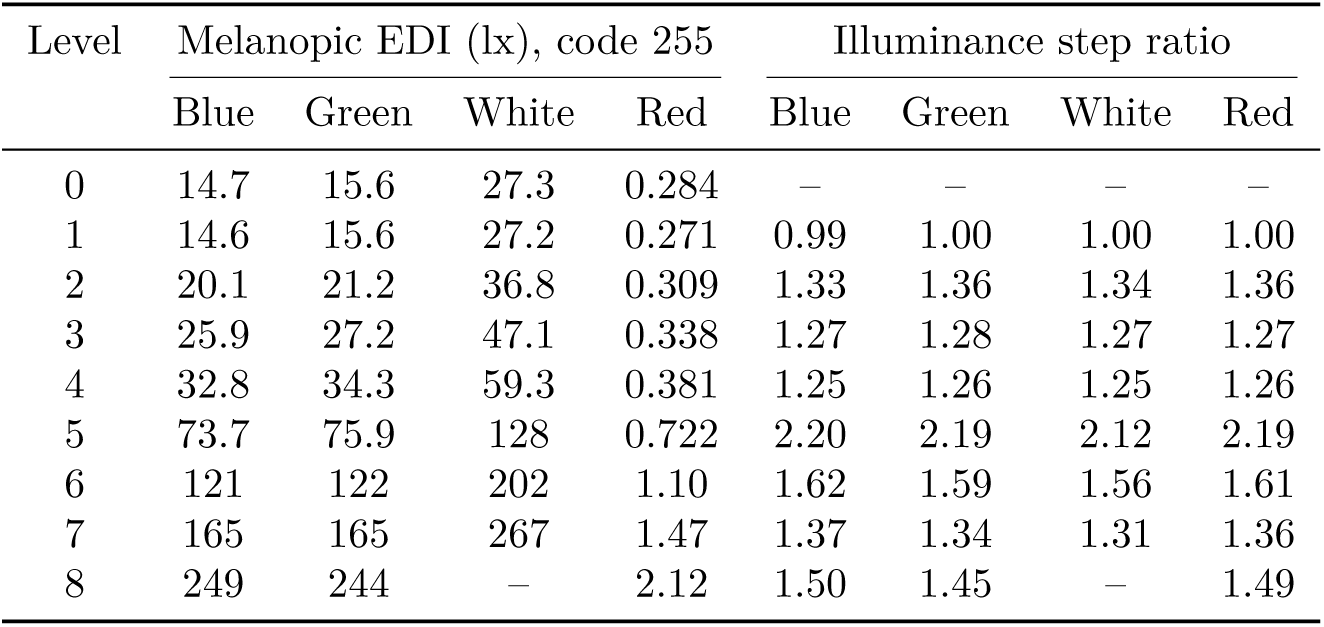
Hardware brightness lookup for each channel at full code (duty cycle 98; means of three sessions). Step ratios are the photopic illuminance of each level divided by that of the level below. White is limited to level 7.

**Table 12:**
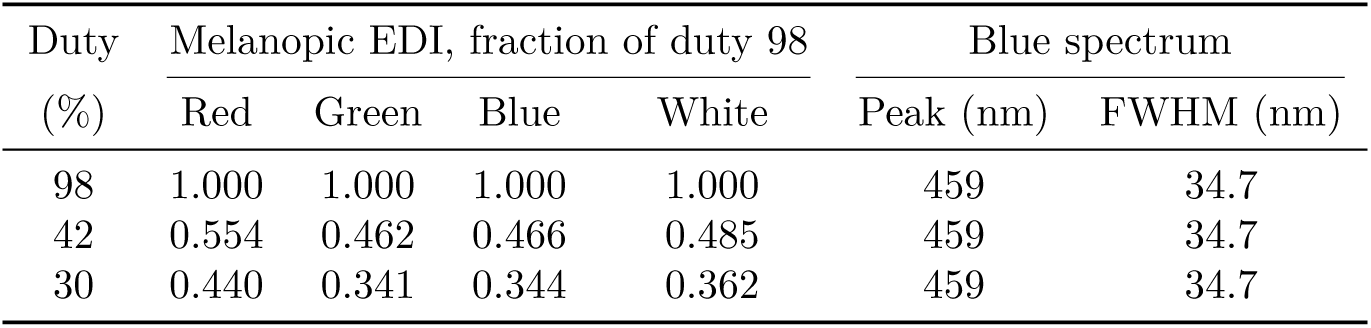
Duty-cycle series: melanopic EDI of each channel at full code (red, green, and blue at level 8, white at level 7) as a fraction of its value at duty 98, and the blue spectrum at each duty (means of three sessions).

| Duty (%) | Melanopic EDI, fraction of duty 98 |  |  |  | Blue spectrum |  |
| --- | --- | --- | --- | --- | --- | --- |
|  | Red | Green | Blue | White | Peak (nm) | FWHM (nm) |
| 98 | 1.000 | 1.000 | 1.000 | 1.000 | 459 | 34.7 |
| 42 | 0.554 | 0.462 | 0.466 | 0.485 | 459 | 34.7 |
| 30 | 0.440 | 0.341 | 0.344 | 0.362 | 459 | 34.7 |

**Table 13:** Example application frames: exposure predicted from the locked model, at the frame’s level and duty cycle 98, and measured (mean of 3 sessions). Predicted: the pixel-weighted sum of uniform predictions (equal spatial weighting). Mean RGB: the frame predicted as a uniform field of its mean code. Spatial: the spatial correction model, with the grid’s weight map and lit-area gain, applied without fitting to the frame measurements (Section 2.6). Reference sum: the same-session uniform references of the frame’s colors, weighted by pixel fraction; spatial reference sum: the same references with the spatial weighting. Measured values are net of black.

|  | Morning text | Blue orb | Night text | Red orb |
| --- | --- | --- | --- | --- |
| Hardware level $L$ | 8 | 8 | 5 | 5 |
| Non-black pixels (%) | 12 | 44 | 100 | 44 |
| *Predicted* |  |  |  |  |
| Melanopic EDI (lx) | 31.2 | 52.6 | 0.663 | 0.106 |
| Photopic illuminance (lx) | 3.72 | 6.31 | 5.05 | 5.82 |
| Melanopic DER | 8.37 | 8.33 | 0.131 | 0.0183 |
| Melanopic EDI, mean RGB (lx) | 3.82 | 19.7 | 0.115 | 0.0272 |
| Melanopic EDI, spatial (lx) | 47.8 | 80.2 | 1.01 | 0.169 |
| Photopic illuminance, spatial (lx) | 5.58 | 9.50 | 7.44 | 9.08 |
| *Measured* |  |  |  |  |
| Melanopic EDI (lx) | 47.9 | 80.8 | 0.874 | 0.166 |
| Photopic illuminance (lx) | 5.67 | 9.71 | 7.12 | 8.69 |
| Melanopic EDI, reference sum (lx) | 30.3 | 51.9 | 0.566 | 0.109 |
| Melanopic EDI, spatial reference sum (lx) | 46.4 | 78.9 | 0.864 | 0.167 |

### **,A,,.1,** Calibration design and model development

#### Calibration grid

The primaries stage measured every code at the brightest state, and crossed level and duty only where the signal stays well above black; dimmer cells of a full code *×* level *×* duty grid were close to the meter’s photopic minimum and to the variation of their own black captures, and lengthened the session enough for the dark reading to drift. The brightness response at the remaining levels comes from the characterization sessions’ full-code brightness series. The primaries sessions showed that a code below 255 gives a smaller fraction of full-code output at low levels than at the brightest state, so the code response depends on level; and the full-code brightness series rises slowly from level 0 to 4 and steeply from 5 to 8, indicating two dimming regimes. The code-series stage therefore gives each regime its own code series. After a first cross-validation, and before the validation session, the supplemental mixture stage presented the 8 compositions with the largest melanopic error (a dim channel next to a full-code channel, and the low gray axis) at level 6 and duty cycle 98, between the two mixture levels at which that error differed. The two excluded code-series attempts had a blue anchor of 17.1 and 19.8 lx, against 29–31 lx in every other model session of the same day. The validation mixtures at level 5 lie between the mixtures’ calibrated levels, so the interaction term is interpolated rather than extrapolated in level (level 5 is also a code-series level of the primaries), and those at level 6 share the supplemental stage’s level but not its compositions. Block order was randomized with a recorded seed and reversed in alternate sessions, and the full-code blue anchor was captured between blocks; a failed anchor was retaken once, and two consecutive anchor failures ended the session.

#### Model terms

The code response *a_c_* is a free parameter (a code node) at every code below 255 and code-series level at which that code was measured (levels 2, 4, 5, 7, and 8), because a single power law per level fitted level 8 within 2% but missed code 64 at levels 2 and 4 by more than half. Between code nodes, *a_c_* follows power-law segments in code and is interpolated log-linearly in level between code-series levels; below a level’s lowest node code, the lowest segment is extended toward code 0, an extrapolation that the estimator flags (Step 2). Codes below 255 are not supported below level 2. *q_c_* is interpolated linearly between calibrated duty cycles without extrapolation (Step 3), and *b_c_* is estimated jointly at every hardware level, the levels outside the model grid from the full-code brightness series of the characterization sessions. A crossed form instead interpolates code and duty within the measured grid at each grid level and transfers to other levels by the ratio *b_c_*(*L*)*/b_c_*(*L*^∗^) to the nearest grid level *L*^∗^ (Crossed models, above). Both interaction forms use *g* = *β*_0_ + *β_L_L/*8 + *β_D_D/*100, fitted to the mixtures with weak ridge regularization (*λ* = 10^−3^); the total form multiplies the sum by 1 *− E g* with *E* the drive in excess of the strongest channel, and the per-channel form attenuates each primary by the drive *O_c_* of the other channels (Step 5). The per-channel form was added after the calibration mixtures showed that a dim channel next to a full-code channel loses output while the full-code channel does not (blue at code 64 with full red at level 7 delivered about two thirds of its own melanopic EDI, while the photopic sum, carried by red, was unchanged); it was selected or rejected by the same cross-validation as the other candidates.

#### Red melanopic output from its DER

Red’s melanopic EDI is small, so at levels 2 and 4 its dim codes fall below the melanopic scoring floor, and a cross-validation fold can lack the red melanopic code nodes that mixtures containing dim red need. The candidate with red’s melanopic output from its DER therefore uses red’s photopic code and duty responses for melanopic EDI, with the full-code level values multiplied by red’s melanopic DER, the geometric mean of melanopic EDI over photopic illuminance across red’s single-channel settings eligible for both outputs (0.018; 10 settings, all within -3% to +5% of it). This assumes that red’s spectral shape, and so its DER, does not change with code, level, or duty, as the measured settings indicate.

#### Candidates

The seven candidates are the factorized and crossed forms, each with no interaction term, the total form, or the per-channel form, and the factorized per-channel form with red’s melanopic output from its DER (Table 6).

#### Cross-validation results

Cross-validation on the calibration sessions gave a leave-one-session-out MAPE of 6.1% for melanopic EDI and 4.9% for photopic illuminance for the selected model, with no failed predictions; the same model without red’s melanopic output from its DER failed 33 melanopic predictions (Table 6). By group, the cross-validation MAPE was 2.9% (melanopic EDI) and 4.4% (photopic) for single primaries and 8.0% and 5.4% for mixtures (95th percentile 22.1% for mixture melanopic EDI), so the largest error was the mixtures’ melanopic EDI; it was predicted better in validation, where the mixtures lay at levels between or at the calibrated mixture levels. With every session scaled by its blue anchor, the MAPE was 5.5% and 3.9% (7.4% for mixture melanopic EDI); the difference is the between-session scale left by reseating, which a wearer also experiences.

#### Selection details

Each cross-validation fold refits every parameter with the characterization brightness series as a fixed input; the worst-case error is the larger of the two outputs’ MAPE. The fitting software first prefers the simplest candidate within its default model-development thresholds (MAPE *≤* 5% and P95 *≤* 10% for both outputs, with no failures); no candidate met them, so the candidate with the fewest failed predictions and then the lowest worst-case MAPE was selected. These thresholds are engineering preferences of the software, not a prespecified criterion for declaring the model successful. As a secondary analysis, the cross-validation was repeated with each calibration session scaled by the mean blue anchor of all sessions divided by its own, which removes the between-session scale left by reseating the glasses; it is reported alongside the primary result and was not used for selection, because a wearer has no anchor measurement. Because codes below 128 were measured only at the highest duty cycle, the crossed forms are supported only near their grid nodes, and whether duty scaling depends on code or level is tested only at codes 128 and 255 and levels 4 and 8; the comparison therefore mainly checks the factorized form rather than offering a general alternative.

#### Model-domain caveats

Below a level’s lowest code node, predictions are extrapolated and flagged, and at levels 2 and 4 the extrapolation underestimated dim codes in the calibration mixtures. Multi-channel stimuli are supported only at levels 4–7. Red’s melanopic output assumes a constant red DER. In cross-validation, mixture melanopic EDI had a larger prediction error than single primaries, because the loss of a dim channel next to a bright one changes with level and the calibrated mixture levels resolve that change only coarsely. For the spatial correction model, the lit-area gain rests on four grid points at one level and duty cycle. The four static frames it matched, at mean drives of 0.12–0.21, were measured before the correction was formulated and do not constitute an independent validation.

#### Eligibility

A setting was eligible for an output if its gross photopic illuminance was at least 1 lx (a photopic criterion, applied to both outputs), and the output’s display-attributable value was positive and at least 3 times a background-variation proxy: the larger of the change between the two black captures bracketing its block and 10% of their mean. This proxy is a conservative margin over the observed black variation, not an estimated noise standard deviation, so the criterion is not a statistical significance test. The photopic minimum alone does not ensure a usable melanopic signal, so the melanopic reading of a setting can be excluded while its photopic reading is kept. For scoring, the melanopic EDI also had to be at least 0.5 lx: red alone read 0.1–0.3 lx melanopic EDI at levels 0–4 whatever its code, a floor of the meter rather than display output. The same rule without this floor selected the settings used for fitting, so the log-space fits use only positive targets, and a channel whose own melanopic output is at the floor still has parameters for the mixtures it is part of. An eligible setting whose predicted photopic illuminance is below 1 lx stays in the error summary and is counted. DER errors use the settings eligible for both outputs with both predictions available. The mean absolute error (MAE, lx) of an output is computed over every quantitative validation setting for which the model returned that output, including settings not eligible for it, because percentage errors of dim settings are large for small absolute differences and because excluding dim settings could hide absolute bias there.

#### Spatial correction model

A spatial weight map, a two-dimensional Gaussian on a constant normalized to mean 1 over the frame, was fitted to the mean photopic cell weighting of red, green, and blue in the grid sessions. A lit-area gain was modelled as a power law in the frame’s mean drive, the pixel mean of the summed photopic code responses (1 for a full-field primary, 3 for full white), *G* = load^−^*^α^*, with *α* fitted in log space through the primaries’ mean cell sum (load 1/9), the full field (load 1), full white over the sum of its primaries (load 3), and the white cells (load 1/3), separately for melanopic EDI and photopic illuminance (*α* = 0.079 photopic and 0.090 melanopic EDI). Each color’s uniform prediction was then weighted by the mean of the weight map over its pixels and scaled by the ratio of the frame’s gain to the color’s own full-field gain; the same weighting applied to the same-session references gives a spatially corrected reference sum.

## B Lookup tables

The brightness and duty-cycle lookups of the characterization sessions and the numerical results of the example application frames, summarized graphically in Figs. 3 and 5. Software should take these values from the calibration package rather than from the tables.

## C Platform, acquisition, and photometric details

### Platform

The platform configuration is listed in Table 14. The application can set the left and right halves of the framebuffer independently; in the display mode used here both eyes see the same full frame, so both halves were set to the same stimulus for every measurement, and readback covers both halves. Whether the 1920*×*1080 frame is scaled or letterboxed onto the 1920*×*1200 panels was not determined. The glasses’ Wearer Detection feature, which turns the display off one minute after the glasses are removed, cannot be read or changed through the SDK, so it was disabled for all measurements on the phantom. Each event-log record is a timestamped JSON object that also contains the eye target of the stimulus. Measurement software drives the application over an opt-in localhost control port; each command is answered only after the next frame has been presented, with the full logged state and a framebuffer readback.

**Table 14:** Platform configuration of the measurements.

| Item | Setting |
| --- | --- |
| Glasses | VITURE Luma Ultra, firmware 12.0.01.101_20260605, film off |
| Control library | VITURE Glasses SDK 2.4.0 (USB) |
| Display mode | 0x31 (2D, 1920×1080 at 60 Hz, same frame to both eyes) |
| Host | Samsung 930XED (12th Gen Intel Core i5, 8 GB RAM), Microsoft Windows 11 |
| Host application | `chronolume_host`, C++17 with GLFW/OpenGL, commit 88fb6cf |
| Framebuffer | 1920×1080, extended (not mirrored) display |
| Renderer | GL_RGB8 textures, nearest-neighbor sampling |
| Color processing | sRGB framebuffer conversion and HiDPI scaling off in the application;<br>HDR, Night light, and automatic color management off in Windows |
| Duty cycle | 98 (held except in the duty series); presets 30, 42, and 98<br>Initial state 90 (not a preset), read at session start |
| Wearer Detection | disabled with the manufacturer’s web firmware tool [30] |

### Characterization sequence

The three sessions were run on 4 October 2026. Each session comprised the same 62 captures of 3 repeats each: full-code red, green, blue, and white; blue codes (0, 0*, B*) for *B ∈ {*16, 32, 64, 128, 192, 255*}*, taken from brightest to dimmest and followed by black; red, green, and white codes 64, 128, and 192; each channel at full code at every hardware brightness level, from level 8 down to level 0; and a duty series of full-code red, green, and blue at level 8 and white at level 7 at the other preset duty cycles, 42 and 30. Duty was changed only while the display was black. Steps without their own brightness level ran at level 8 and steps without their own duty cycle at duty 98; because full white was limited to level 7, white steps at the base level ran at level 7 and the white brightness series spans levels 0–7.

### Example frames

Exact frame definitions are in Table 15. Each frame was a block at its own level and the base duty cycle: black, the frame, each of its non-black colors as a uniform full-field reference, and black again, followed by the full-code blue anchor. The frame order was reversed in alternate sessions.

**Table 15:** Example application frames as rendered (RGB codes).

| Frame | Content | Non-black pixels (%) | Level |
| --- | --- | --- | --- |
| Morning text | bold (0, 0, 255) text on black | 12 | 8 |
| Blue orb | disc the height of the frame, concentric rings at blue<br>codes 255, 192, 128, 64 from the center out, on black | 44 | 8 |
| Night text | (255, 92, 56) text on a (20, 5, 7) background | 100 | 5 |
| Red orb | the same orb in red, on black | 44 | 5 |

### Full white at level 8

In preliminary sweeps, showing (255, 255, 255) at level 8 and duty cycle 98 collapsed the display output and then dropped the glasses’ USB control link until the host application was restarted, even when brightness had been set with the display black. The failure was reproduced on a macOS host at duty cycle 90, where raising white from level 7 to 8 was refused by the SDK or the display dropped out, so it is a property of the device rather than of the host platform. Red, green, and blue alone ran normally at level 8, and white ran normally at level 7. The measurement software therefore never presents white above level 7 and records the limit with each affected capture.

### Quality-control criteria

Each capture was subject to the following checks, all computed from its own readings:

,•, *Readback.* Before every capture, the application framebuffer readback of both halves must match the requested code on a full-size framebuffer.
,•, *No display light.* The primaries peak at 459, 539, and 631 nm, so a reading of a non-black stimulus whose spectrum peaks at or above 700 nm is the meter’s dark noise with no display light. That reading is discarded and retaken.
,•, *Repeat outliers.* If any repeat differs from the capture median by more than 30% (checked when the median is at least 0.1 lx), the whole capture is re-run, up to three times.
,•, *Anchors.* An anchor more than 10% from the first of its session stops the sweep and re-runs the preceding block, up to twice.
,•, *Unit check.* Photopic illuminance computed from the exported spectrum must match the meter’s own illuminance within 3% (checked at *≥*0.5 lx). This confirms the units and integration of the exported spectrum; it is not an independent accuracy check.

### Photometric and α-opic definitions

Photopic illuminance is *E_v_* = *K_m_* ∫*E_λ_V* (*λ*) *dλ* (*K_m_* = 683 lm W^−1^, CIE 1931 2^◦^ *V* (*λ*)). For each retinal photoreceptor class *α* (S-cone-opic through melanopic), the *α*-opic irradiance is *E_α_* = ∫*E_λ_ s_α_*(*λ*) *dλ*, and the equivalent daylight illuminance (EDI) normalizes *E_α_* by the *α*-opic irradiance of standard illuminant D65 at one photopic lux [10]. The melanopic DER is the melanopic EDI divided by the photopic illuminance. From each mean spectrum we also report integrated irradiance (380–780 nm), band irradiance (400–500 nm), photon irradiance (380–780 nm and 400–500 nm), peak wavelength, full width at half maximum (FWHM, by linear interpolation at half maximum), spectral centroid, and the shares of 380–780 nm irradiance in the non-overlapping 380–500, 500–600, and 600–780 nm bands (each band includes its lower and excludes its upper boundary sample, except the last). Anchor stability is the median illuminance of each anchor relative to the first anchor of its session.

### Blue-light-hazard screen

For extended sources the ICNIRP limit applies to weighted radiance averaged over a duration-dependent acceptance angle [27]. The irradiance measured here integrates the whole display, so it cannot be smaller than the weighted irradiance collected within any such acceptance angle; comparing it with the small-source irradiance criterion is therefore conservative with respect to the angular distribution of radiance. For spatial averaging, ICNIRP specifies a 7 mm averaging aperture for inhomogeneous irradiance profiles and permits larger apertures only for homogeneous ones [27]. The active diameter of the diffuser (7 mm) equals the 7 mm aperture, so the measured irradiance is already averaged over the specified aperture and requires no assumption about the homogeneity of the irradiance profile.

